# Sex differences in associations between *APOE ε2* and longitudinal cognitive decline

**DOI:** 10.1101/2022.11.28.22282828

**Authors:** Madeline E. Wood, Lisa Y. Xiong, Yuen Yan Wong, Rachel F. Buckley, Walter Swardfager, Mario Masellis, Andrew S. P. Lim, Emma Nichols, Renaud La Joie, Kaitlin B. Casaletto, Raj G. Kumar, Kristen Dams-O’Connor, Priya Palta, Kristen M. George, Claudia L. Satizabal, Lisa L. Barnes, Julie A. Schneider, Alexa Pichette Binet, Sylvia Villeneuve, Judy Pa, Adam M. Brickman, Sandra E. Black, Jennifer S. Rabin, the ADNI and Prevent-AD Research Groups

## Abstract

**INTRODUCTION:** We examined whether sex modifies the association between *APOE* ε*2* and cognitive decline across two independent samples.

**METHODS:** We used observational data from non-Hispanic White (NHW) and non-Hispanic Black (NHB) cognitively unimpaired adults. Linear mixed models examined interactive associations of *APOE* genotype (ε*2* or ε*4* carrier vs. ε*3/*ε*3*) and sex on cognitive decline in NHW and NHB participants separately.

**RESULTS:** In both Sample 1 (*N*=9,766) and Sample 2 (*N*=915), sex modified the association between *APOE* ε*2* and cognitive decline in NHW participants. Specifically, relative to *APOE* ε*3/*ε*3, APOE* ε*2* protected against cognitive decline in men but not women. Among *APOE* ε*2* carriers, men had slower decline than women. Among *APOE* ε*3/*ε*3* carriers, cognitive trajectories did not differ between sexes. There were no sex-specific associations of *APOE* ε2 with cognitive decline in NHB participants (*N*=2,010).

**DISCUSSION:** In NHW adults, *APOE* ε*2* may selectively protect men against cognitive decline.

## 1. Background

Women have a greater lifetime risk of developing Alzheimer’s disease (AD) dementia than men.^1^ While some studies suggest that women’s increased risk is related to longer survival,^2,3^ other studies report that sex/gender disparities exist beyond what can be explained by female longevity alone.^4^ Mounting evidence suggests that biological mechanisms underpin sex differences in AD risk and progression.^5–10^

The *APOE* gene encodes a protein that transports cholesterol in the blood.^11^ *APOE* ε*3* is the most common allele^12^ and is neutral in relation to risk for AD dementia.^11^ *APOE* ε*4* is associated with a higher risk of AD dementia^13^ (mostly in non-Hispanic White populations^14^), whereas *APOE* ε*2* is associated with a lower risk of AD dementia.^15^ Accumulating data suggest that there are sex differences in the effects of *APOE* ε*4* on AD risk. Studies report that women who carry *APOE* ε*4* are disproportionately vulnerable to cognitive impairment^16^ and AD^15^ compared to their counterpart men.

Although a less robust literature, *APOE* ε*2* may also have sex-specific effects on AD risk. The few reports on sex-specific effects of *APOE* ε*2* have been in the context of studies focused on *APOE* ε*4* sex differences. One study found that in men but not women, *APOE* ε*2* carriage was associated with reduced risk of progression from normal cognition to mild cognitive impairment (MCI) or AD dementia.^16^ By contrast, a meta-analysis found that in cognitively unimpaired older adults, *APOE ε2*/*ε3* carriage decreased the risk of AD dementia more strongly in women than in men.^17^ That same meta-analysis reported the opposite pattern for *APOE ε2* homozygosity (<30/sex), such that *APOE ε2*/*ε*2 carriage was protective against AD dementia in men but not in women.^17^ Other studies examining sex-specific effects of *APOE ε2* on cognition have also yielded mixed results, with some showing greater protection for women and others showing greater protection for men.^18–20^ These studies had small numbers of *APOE ε2* carriers, and were cross-sectional in design or had limited longitudinal follow-up.^18–20^

Allele frequencies can vary widely between populations of different ancestral backgrounds (i.e., population stratification), which can lead to unreliable associations between genetic factors and phenotypic outcomes.^21–23^ There is evidence that *APOE ε4* confers differential risk for AD across races. While *APOE ε4* carriage is more common among Black (vs. White) populations, the association of *APOE ε4* with risk for cognitive decline and AD dementia appears to be attenuated in Black adults.^24–26^

In the present study, we carried out an in-depth investigation of sex differences in associations between *APOE ε2* carriage and longitudinal cognition. We first examined sex differences using pooled data from cognitively unimpaired adults participating in either the National Alzheimer’s Coordinating Center (NACC) or Rush Alzheimer’s Disease Center cohort studies (Sample 1). To control for population stratification^21–23^ and potentially differing effects of *APOE* across racial/ethnic groups,^24–26^ we performed analyses separately in non-Hispanic White (NHW) and non-Hispanic Black (NHB) participants. On finding sex-specific effects in NHW participants, we then sought to replicate the main findings in an independent sample of participants from Alzheimer’s Disease Neuroimaging Initiative (ADNI) and Pre-symptomatic Evaluation of Experimental or Novel Treatments for Alzheimer Disease (Prevent-AD) (Sample 2).

## 2. Methods

### 2.1. Participants

Data were obtained from four independent sources: 1) NACC; 2) Rush Alzheimer’s Disease Center cohort studies: Religious Orders Study (ROS), Rush Memory Aging Project (MAP), and Minority Aging Research Study (MARS); 3) ADNI; and 4) Prevent-AD. Sample 1 consisted of data from NACC and ROS/MAP/MARS. Sample 2 consisted of data from ADNI and Prevent-AD. Research procedures were approved by the relevant ethics committees and participants provided written informed consent. This study follows the Strengthening the Reporting of Observational Studies in Epidemiology (STROBE) reporting guidelines for cohort studies.

Since *APOE ε2* protects against cognitive decline,^27^ we restricted our sample to participants classified as cognitively unimpaired at baseline. This allowed us to maintain a representative proportion of *ε2* carriers and to examine early cognitive changes with respect to *APOE* genotype. We also required that participants were ≥50 years old at baseline and had at least one follow-up cognitive assessment. In NACC, cognitively unimpaired is defined as a Clinical Dementia Rating (CDR) global score of 0.^28^ In ROS/MAP/MARS, cognitively unimpaired is defined as the absence of MCI or dementia.^29,30^ In ADNI and Prevent-AD, cognitively unimpaired is defined according to several criteria, one of which is a CDR global score of 0.^31,32^ In the present study, we only included participants who self-identified as NHW and NHB, since these were the largest racial/ethnic groups across data sources (see supplemental material for details on how race and ethnicity was coded). Details on the sample selection processes are described in Figure S1 in the supplemental material.

### 2.2. Cognition

All four data sources (i.e., NACC, ROS/MAP/MARS, ADNI, Prevent-AD) assess cognition approximately annually. Across the data sources, we created a comparable cognitive composite that was weighted towards episodic memory (see supplemental material for specific tests). To calculate the composite, we z-transformed the raw test scores using the mean and standard deviation of the baseline study samples, and then computed the average of the standardized scores.

### 2.3. Genotype

We used publicly available *APOE* genotype data to classify participants as *ε2, ε3/ε3*, or *ε4* carriers. Our samples had relatively few *APOE ε2* homozygotes (*N*=56 in NACC; *N*=13 in ROS/MAP/MARS, *N*=1 in ADNI, *N*=0 in Prevent-AD), and therefore participants with one or two copies of *ε2* were categorized as *ε2* carriers. Participants with one or two copies of *APOE ε4* were categorized as *ε4* carriers. *APOE ε3* homozygotes were the reference group. *APOE ε2/ε4* carriers were excluded due to the opposing effects of *ε2* and *ε4* alleles on AD risk.^27^ All samples met Hardy-Weinberg Equilibrium expectations.^33^

### 2.4. Statistical Analysis

Analyses were conducted in R (v.4.1.2). We used t-tests and χ^2^ tests to assess differences in demographic and baseline characteristics between men and women. We used linear mixed models to examine the interactive effects of *APOE* allele (*ε2* and *ε4* vs. reference ε*3/*ε*3*), sex (reference female), and time (years from baseline) on longitudinal cognition separately in NHW and NHB participants. Where possible sex-specific *APOE ε2* effects were observed, we then performed sex- and genotype stratified analyses. Sex-stratified analyses examined the two-way interaction between *APOE* allele and time on cognition, allowing us to compare *APOE ε2* versus *ε3/ε3* cognitive trajectories in men and women separately. Genotype-stratified analyses examined the two-way interaction between sex and time on cognition, allowing us to compare cognitive trajectories of men and women *APOE ε2* carriers as well as men and women *APOE ε3/ε3* carriers. All models included random intercepts and slopes. As in a previous study,^34^ including an additional quadratic term for time (to account for accelerated decline with aging) resulted in better model fit compared to models without this term (*p*<.05). Therefore, all models included this term.

We first examined sex differences in associations between *APOE ε2* and cognitive decline in NHW and NHB participants from Sample 1. On finding sex-specific effects in NHW participants, we sought to replicate these effects in an independent sample of NHW participants (Sample 2). In exploratory analyses, we examined whether the sex-specific effects of *APOE ε2* on longitudinal cognition were more pronounced at older ages. To do so, we repeated the main analyses after restricting the baseline age according to four cut-off values: age ≥65, ≥70, ≥75, and ≥80 years. Finally, to contextualize our findings, effect sizes for sex-specific *APOE ε2* findings were compared against sex-specific *APOE ε4* findings.^15,16^

#### 2.4.1 Covariates

In all analyses, we adjusted for data source (i.e., NACC vs. ROS/MAP/MARS or ADNI vs. Prevent-AD), baseline age, years of education, and their interactions with time. To account for practice effects on neuropsychological tests, we included a term for the square root of the number of previous study visits (assumes the largest improvement in performance after the first testing session, with diminishing returns on subsequent sessions).^35^ If this covariate was not significant, it was removed from the models. Because vascular risk factors are associated with cognitive decline,^36,37^ we also adjusted for baseline vascular risk and its interaction with time. Vascular risk was quantified using a summary score^38^ that includes the presence/absence of up to five conditions (diabetes, hypertension, high cholesterol, stroke, and heart conditions; see supplemental material for details).

## 3. Results

### 3.1 Demographic characteristics

Table 1 summarizes the demographic characteristics for NHW and NHB participants in Sample 1 and NHW participants in Sample 2 (Tables S1 and S2 summarize demographic data for each data source separately). In Sample 1 (NACC and ROS/MAP/MARS), 9,766 NHW and 2,010 NHB participants met inclusion criteria. In Sample 2 (ADNI and Prevent-AD), 915 NHW participants met inclusion criteria. With respect to NHW participants, Sample 1 was slightly older than Sample 2 (73.0 years vs. 70.1 years), had a higher proportion of women (65.0% vs 59.1%), a slightly higher proportion of *APOE ε2* carriers (12.9% vs. 11.8%), and more longitudinal follow up (median 6 vs. 5 visits).

**Table 1.**
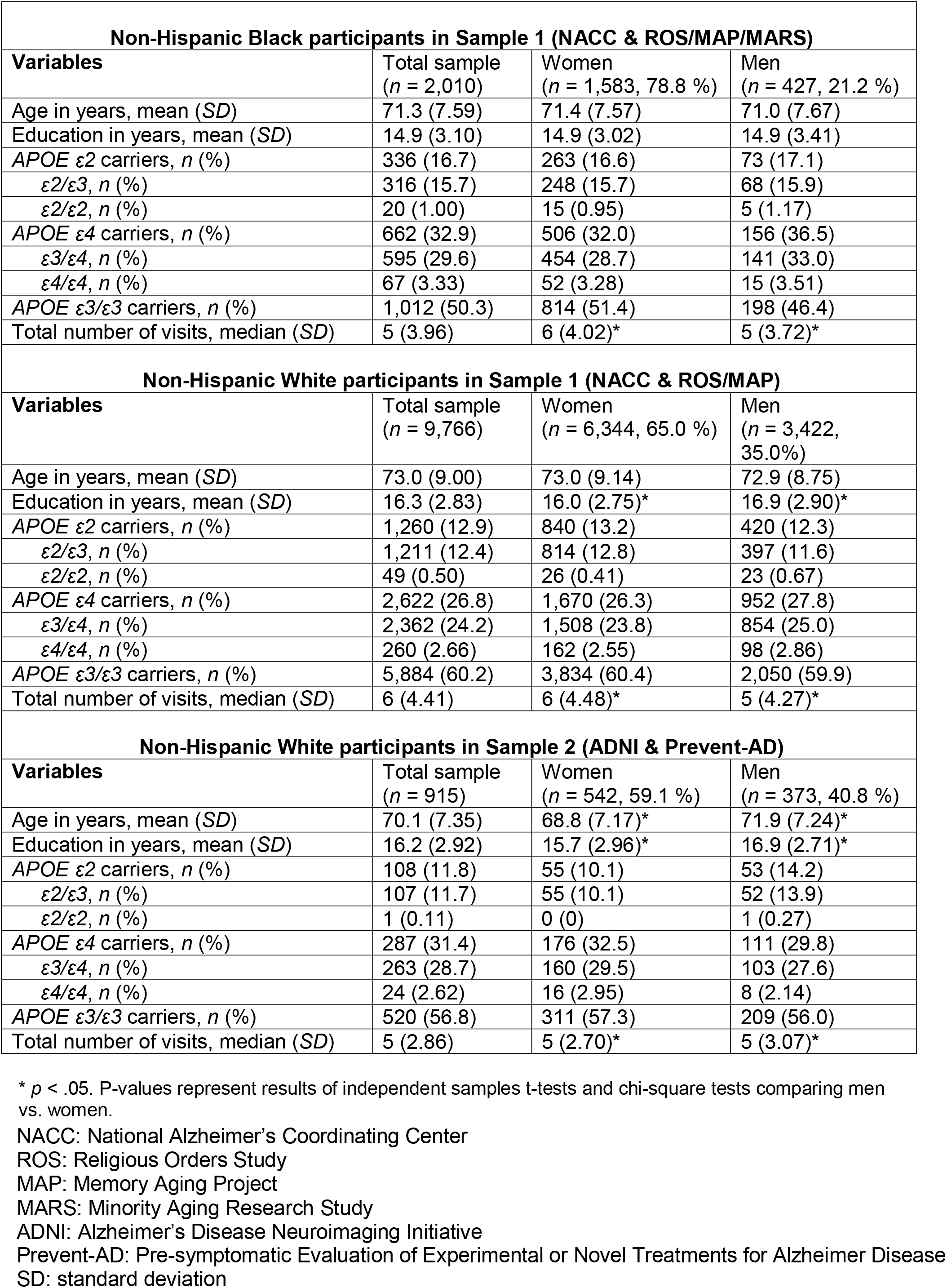
Baseline demographic and clinical characteristics by racial/ethnic group and cohort.

### 3.2 Sex-specific associations of *APOE ε2* with cognitive decline in NHB participants

In Sample 1, the interaction between sex, *APOE ε2*, and time on cognitive decline was not significant in NHB participants (*β*= −0.011, 95% CI: −0.153–0.131, *p*=.88; Table S3; Figure 1). Because there was no evidence of sex-specific *APOE ε2* effects, we tested the two-way interaction between *APOE ε2* (vs. *ε3/ε3*) and time on cognitive decline (adjusting for sex). We found a trend towards slower decline among *APOE ε2* carriers after adjusting for sex (*β*=0.046, 95% CI: −0.012–0.104, *p*=.12; Table S3; Figure S2). With respect to *APOE ε4*, we observed a near significant interaction between male sex, *APOE ε4*, and time in NHB participants (*β*=0.103, 95% CI: −0.017–0.223, *p*=.09; Table S3; Figure S3). Sex- and genotype-stratified analyses showed that women carrying *APOE ε4* exhibited faster cognitive decline relative to both women carrying ε*3/*ε*3* and men carrying *ε4* (Table S3).

**Figure 1.**
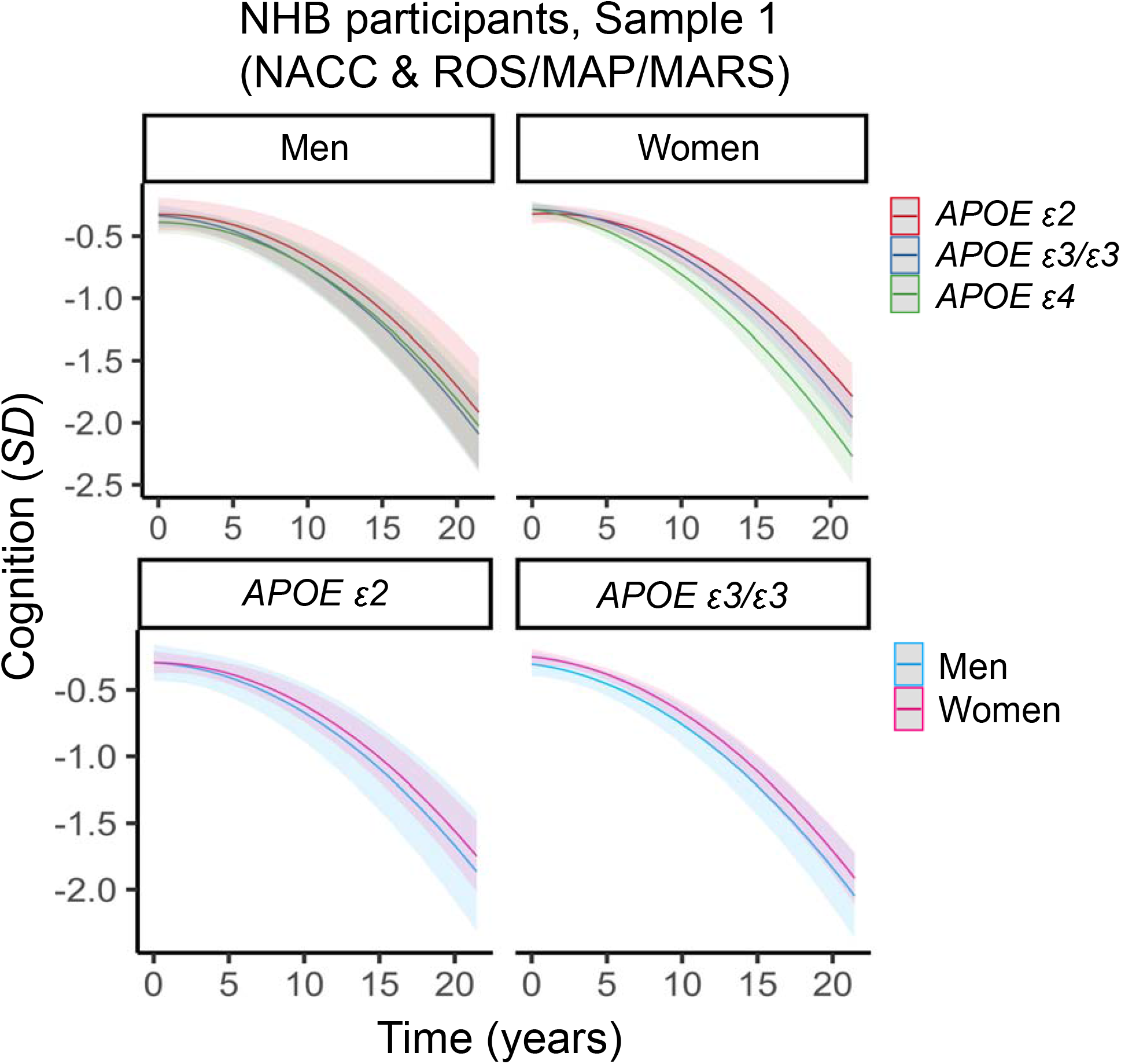
Three-way interaction between sex, *APOE* and time on cognitive decline in non-Hispanic Black (NHB) participants in Sample 1 (NACC & ROS/MAP/MARS). Plots depict marginal effects, showing change in cognition (standardized score) over time, stratified by sex and genotype (*APOE ε4* plot not shown). There were no significant sex differences in associations between *APOE ε2* and global cognitive decline. The models are adjusted for baseline age, years of education, and vascular risk, and their interactions with time. Shaded regions represent 95% confidence intervals.

### 3.3 Sex-specific associations of *APOE ε2* with cognitive decline in NHW participants

In NHW participants from Sample 1, there was a significant interaction between sex, *APOE ε2*, and time (Table 2; Table S4, Figure 2). In sex-stratified analyses, men carrying *APOE ε2* exhibited slower cognitive decline than men carrying *APOE ε3*/*ε3* (Table 2; Table S4). By contrast, cognitive trajectories did not differ between women carrying *APOE ε2* versus *ε3*/*ε3* (Table 2; Table S4). In genotype-stratified analyses, cognitive trajectories differed by sex among *APOE ε2* carriers, but not among *APOE ε3*/*ε3* carriers. Specifically, among *APOE ε2* carriers, men exhibited slower decline relative to women, whereas rates of decline were similar between men and women carrying *APOE ε3*/*ε3* (Table 2; Table S4).

**Table 2.** Sex-specific associations between *APOE ε2* (vs *APOE ε3/ε3)* and longitudinal cognition in non-Hispanic White participants.

| Analyses | Sample 1 (NACC & ROS/MAP) |  | NACC |  | ROS/MAP |  | Sample 2 (ADNI & Prevent-AD) |  |
| --- | --- | --- | --- | --- | --- | --- | --- | --- |
| | $\beta$ (95% CI) | *p* | $\beta$ (95% CI) | *p* | $\beta$ (95% CI) | *p* | $\beta$ (95% CI) | *p* |
| Three-way interaction: |  |  |  |  |  |  |  |  |
| Sex × *APOE* $\epsilon 2$ (vs. $\epsilon 3/\epsilon 3$) × time | 0.097 (0.004 – 0.031) | .01 | 0.081 (0.010 – 0.152) | .02 | 0.127 (-0.069 – 0.323) | .20 | 0.196 (0.007 – 0.386) | .04 |
| Sex-stratified two-way interactions: |  |  |  |  |  |  |  |  |
| *APOE* $\epsilon 2$ (vs. $\epsilon 3/\epsilon 3$) × time in men | 0.096 (0.037 – 0.155) | .001 | 0.074 (0.020 – 0.128) | .008 | 0.149 (-0.022 – 0.319) | .09 | 0.094 (-0.055 – 0.244) | .22 |
| *APOE* $\epsilon 2$ (vs. $\epsilon 3/\epsilon 3$) × time in women | -0.001 (-0.044 – 0.043) | .97 | -0.008 (-0.051 – 0.035) | .71 | 0.012 (0.089 – 0.114) | .81 | -0.103 (-0.227 – 0.020) | .10 |
| Genotype-stratified two-way interaction: |  |  |  |  |  |  |  |  |
| Male sex × time in *APOE* $\epsilon 2$ carriers | 0.120 (0.051 – 0.190) | .001 | 0.095 (0.028 – 0.161) | .005 | 0.191 (0.012 – 0.371) | .04 | 0.151 (-0.008 – 0.311) | .06 |
| Male sex × time in *APOE* $\epsilon 3/\epsilon 3$ carriers | -0.000 (-0.031 – 0.030) | .99 | -0.005 (-0.033 – 0.024) | .75 | 0.038 (-0.044 – 0.121) | .36 | -0.005 (-0.088 – 0.077) | .90 |

NACC: National Alzheimer's Coordinating Center
ROS: Religious Orders Study
MAP: Memory Aging Project
ADNI: Alzheimer's Disease Neuroimaging Initiative
Prevent-AD: Pre-symptomatic Evaluation of Experimental or Novel Treatments for Alzheimer Disease
CI: confidence interval

**Figure 2.**
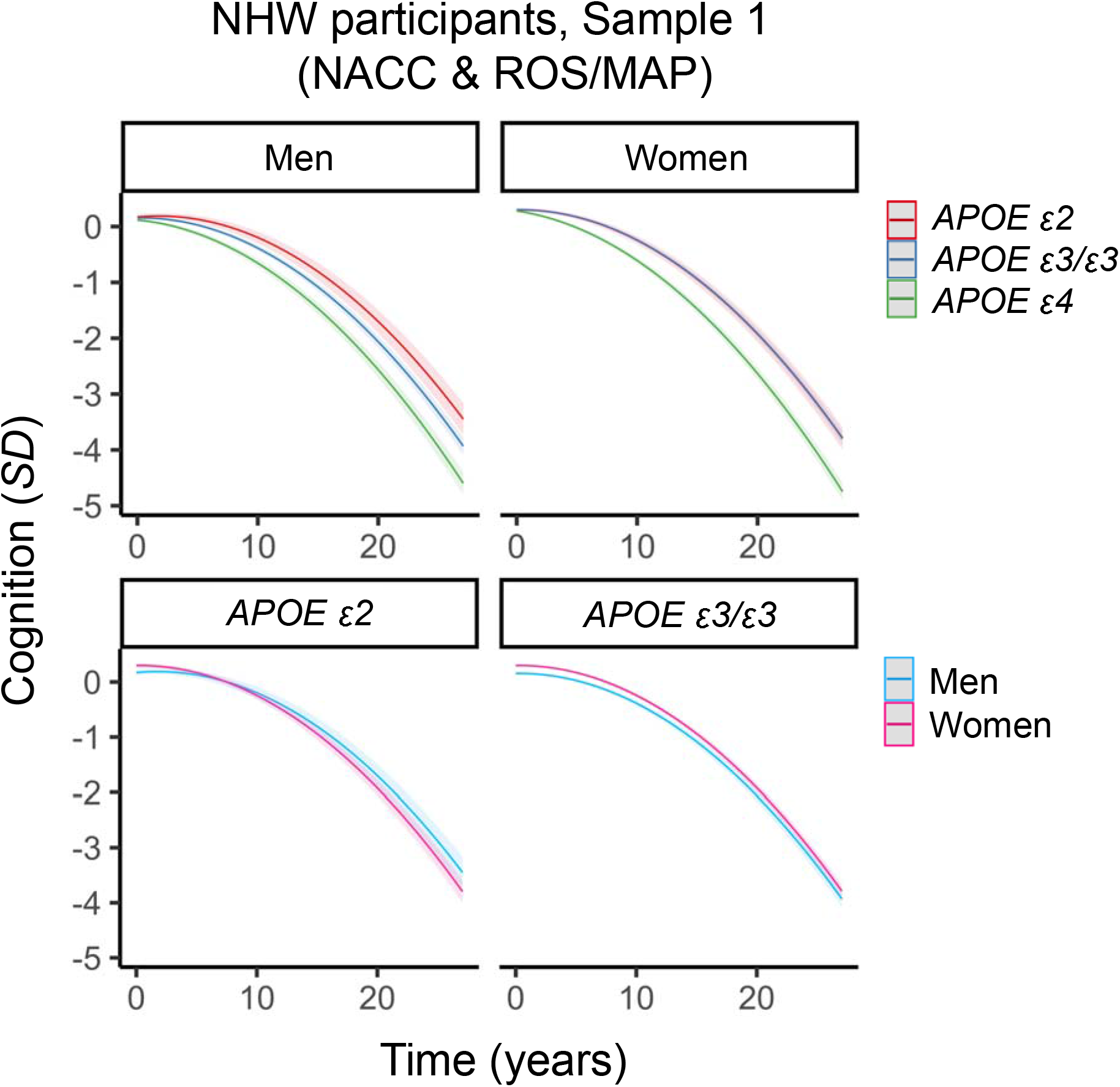
Three-way interaction between sex, *APOE* and time on cognitive decline in non-Hispanic White (NHW) participants in Sample 1 (NACC & ROS/MAP). Plots depict marginal effects, showing change in cognition (standardized score) over time, stratified by sex and genotype (*APOE ε4* plot not shown). In sex-stratified analyses, men carrying *APOE ε2* were more protected against decline than men carrying *APOE ε3*/*ε3*. In women, *APOE ε2* was no more protective than APOE *ε3*/*ε3*. In genotype-stratified analyses, men carrying *APOE ε2* were more protected against decline than women carrying *APOE ε2*. By contrast, rates of decline did not differ between men and women *APOE ε3*/*ε3* carriers. The models are adjusted for baseline age, years of education, and vascular risk, and their interactions with time. Shaded regions represent 95% confidence intervals.

Given the relatively large number of participants in NACC (*N*=7,931, *N* women=4,980, 62.8%) and ROS/MAP (*N*=1,835, *N* women=1,364, 74.3%), we examined whether the pattern of results was present in each data source separately. In NACC, there was a significant interaction between male sex, *APOE ε2*, and time on cognitive decline (Table 2; Table S5, Figure S4, Figure S5). In ROS/MAP, the same three-way interaction was not significant (Table 2; Table S6, Figure S4, Figure S5). However, sex- and genotype-stratified analyses revealed a similar pattern of findings in both data sources (Table 2, Table S5, Table S6). Sex-stratified analyses showed that men carrying *APOE ε2* had a pattern of slower cognitive decline than men carrying *APOE ε3*/*ε3*, whereas women carrying *APOE ε2* did not have slower decline than women carrying *APOE ε3*/*ε3*. In genotype-stratified analyses, men carrying *APOE ε2* had significantly slower decline than women carrying *APOE ε2*. Similarly, men and women *APOE ε3*/*ε3* carriers did not exhibit different cognitive trajectories.

Next, we sought to replicate the main sex-specific findings in an independent sample of NHW participants from ADNI and Prevent-AD (Sample 2). We again observed a significant interaction between male sex, *APOE ε2* and time (Table 2; Table S7, Figure 3). Sex-stratified analyses showed trend-level associations. In men, *APOE ε2* carriers had slower decline than *ε3*/*ε3* carriers, whereas in women, *APOE ε2* carriers had faster decline than *ε3*/*ε3* carriers (Table 2; Table S7). In genotype-stratified analyses, once again, men carrying *APOE ε2* exhibited slower decline than women carrying *ε2*, whereas the rates of decline did not differ between men and women carrying *APOE ε3*/*ε3* (Table 2; Table S7). Importantly given the smaller number of participants in Sample 2, the effect sizes of male specific *APOE ε2* protection in sex- and genotype-stratified analyses were equivalent to or larger than those observed in Sample 1 (i.e., *APOE ε2* (vs. *ε3*/*ε3*) × time in men: Sample 2 *β*=0.094, *p*=.22 vs. Sample 1 *β*=0.096, *p*=.001; Male sex × time in *APOE ε2* carriers: Sample 2 *β*=0.151, *p*=.06 vs. Sample 1 *β*=0.120, *p*=.001).

**Figure 3.**
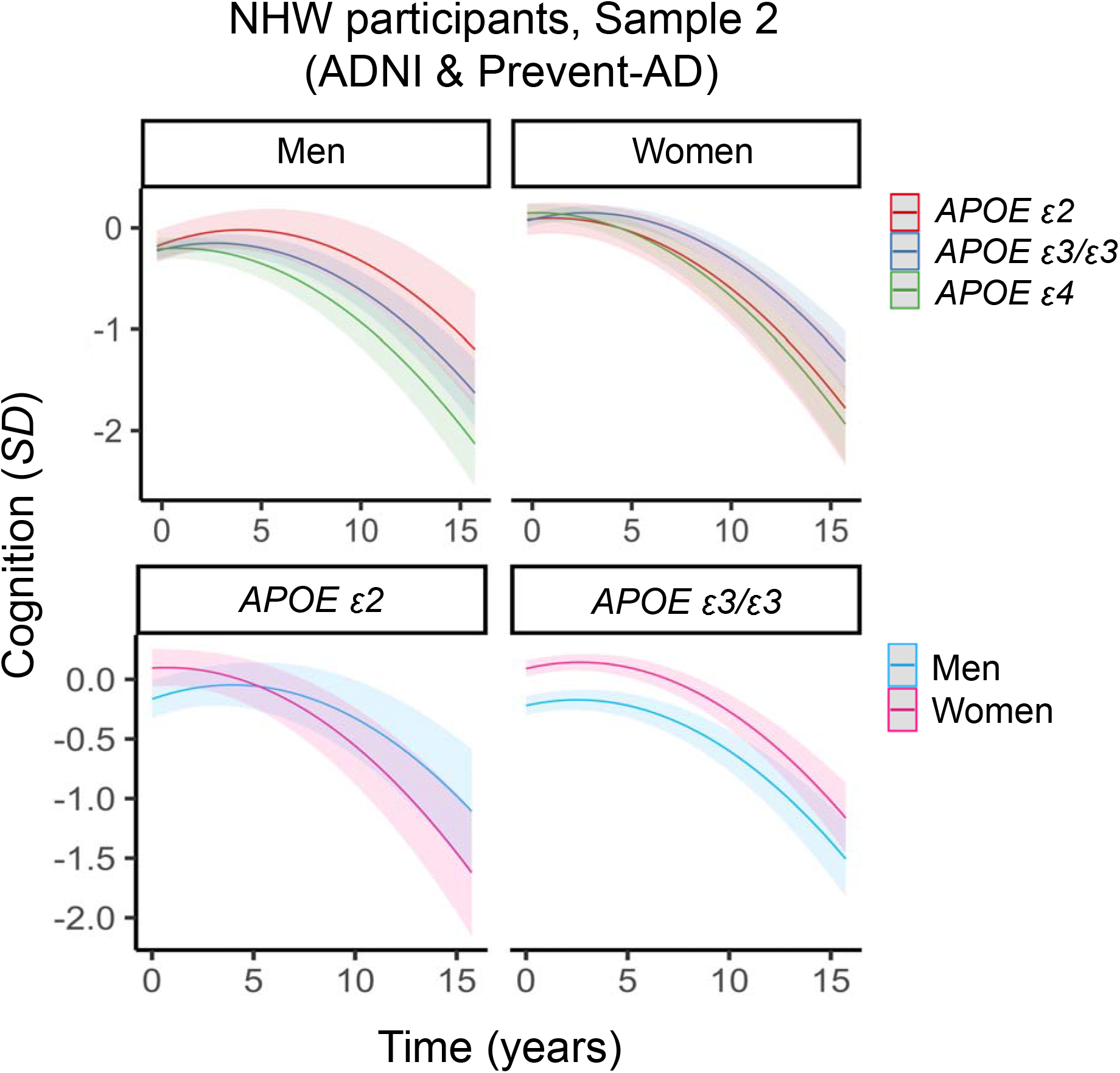
Three-way interaction between sex, *APOE* and time on cognitive decline in non-Hispanic White (NHW) participants in Sample 2 (ADNI & Prevent-AD). Plots depict marginal effects showing change in cognition (standardized score) over time, stratified by sex and genotype (*APOE ε4* plot not shown). Sex-stratified analyses showed trend-level associations. In men, *APOE ε2* was more protective against decline than *APOE ε3*/*ε3*, whereas in women, *APOE ε2* was less protective than *APOE ε3*/*ε3*. In genotype-stratified analyses, men carrying *APOE ε2* were more protected against decline than women carrying APOE *ε2*, whereas the rates of decline did not differ between men and women carrying *APOE ε3*/*ε3*. The models are adjusted for baseline age, years of education, and vascular risk, and their interactions with time. Shaded regions represent 95% confidence intervals.

In exploratory analyses, we examined whether the sex-specific effect of *APOE ε2* on cognitive decline differed across increasing baseline age cut-offs (age ≥65, ≥70, ≥75, and ≥80 years). In Sample 1, we observed that the magnitude of the 3-way interaction term increased as baseline age increased (Table S8). In Sample 2, we observed a similar pattern of increased magnitude among ages 50 through 70 (Table S9). However, the magnitude of the interaction term began to decrease above the age of 75. This is likely due to the considerably smaller sample sizes at these older ages (Table S9). Together, these findings suggest that male-specific *APOE ε2* protection may become more pronounced in older age.

### 3.4 Sex-specific associations of *APOE ε4* with cognitive decline in NHW participants

To contextualize the *APOE ε2* findings in NHW participants in Sample 1 and Sample 2, we sought to replicate previously reported sex differences in associations between *APOE ε4* and cognitive decline. In Sample 1, there was a significant interaction between male sex, *APOE ε4*, and time on cognition in NHW participants (*β*=0.064, 95% CI: 0.007–0.120, *p*=.03; Table S4; Figure 2). Sex stratified analyses demonstrated that *APOE ε4* (vs. *APOE* ε*3*ε*3*) was more strongly associated with cognitive decline in women than men. Genotype stratified analyses showed that women carrying *APOE ε4* declined faster than men carrying *APOE ε4* (Table S4; Figure S6). These same findings were not observed in Sample 2, as the interaction between male sex, *APOE ε4*, and time on longitudinal cognition was not significant (*β*=0.041, 95% CI: −0.095–0.177, *p*=.56; Table S7; Figure 2).

## 4. Discussion

Across two independent samples of cognitively unimpaired NHW participants (Sample 1: NACC and ROS/MAP, Sample 2: ADNI and Prevent-AD), we found that men carrying *APOE ε2* were more protected against cognitive decline compared to both men carrying *APOE ε3*/*ε3* and women carrying *APOE ε2*. Notably, no sex differences were observed among *APOE ε3*/*ε3* carriers. Analyses performed separately in NACC and ROS/MAP showed the same pattern of male-specific protection in *APOE ε2* carriers. In both Sample 1 and Sample 2, the magnitude of the sex-specific *APOE ε2* effect was generally more pronounced at older ages when risk for AD is higher.^39^ We did not observe sex-specific associations in NHB participants. The replication of these findings in cognitively unimpaired NHW adults across each NACC, ROS/MAP and ADNI/Prevent-AD provide compelling evidence that *APOE ε2* protects men but not women against cognitive decline.

The biological mechanisms driving the observed sex differences in the NHW participants are unclear. One possibility may relate to sex hormones, which regulate ApoE protein synthesis.^40^ Estrogen upregulates ApoE synthesis,^40,41^ and like other metabolic and neurological systems,^42,43^ estrogen-mediated *APOE* processes may become disrupted around menopause when estrogen levels decline. If so, *APOE ε2* protection against AD pathology and its downstream cognitive effects may be reduced in postmenopausal women. Additional research is needed to elucidate the mechanisms (including the role of hormones) behind sex-specific *APOE ε2* effects.

The finding of sex-specific associations between *APOE ε2* and cognitive decline complements evidence that women (vs. men) carrying *APOE ε4* are at disproportionately higher risk for AD.^15,16,44,45^ We replicated this finding in NHW and NHB from Sample 1 (but not Sample 2), observing that women carrying *APOE ε4* had faster rates of cognitive decline than their counterpart men. Interestingly, in NHW participants from Sample 1, the effect size for the three-way interaction of *APOE ε*2, sex, and time on cognitive decline (*β*=0.097) was greater than that of the equivalent interaction for *APOE ε*4 (*β*=0.064). This suggests that sex-specific protective effects of *APOE ε2* may represent an important yet overlooked contribution to sex disparities in cognitive and AD outcomes.

It is not clear why we did not observe sex-specific associations between *APOE ε2* and longitudinal cognition in NHB participants. Previous research demonstrates that pathological drivers of cognitive decline may differ across races.^46,47^ It is possible that in NHB participants, sex-specific associations of *APOE ε2* with cognition are obscured by more salient predictors of cognitive decline. Alternatively, sex-specific effects of *APOE ε2* may not exist in NHB persons. This idea is consistent with evidence that *APOE* genotypes differentially impact cognition across racial and ethnic groups.^19,24–26,48^ Future work should seek to further clarify these associations in diverse cohorts, particularly in those with neuropathology data.

The major strength of this study is the replication of sex-specific findings across two independent samples of pooled data (and in each NACC & ROS/MAP separately). This is particularly notable given different sampling procedures, demographic characteristics, cognitive tests, and follow-up times across the studies. There are also several limitations. First, study participants are generally well-educated, which may limit the generalizability of our findings. Second, since whole genome sequencing or equivalent data were not available for many study participants, we were unable to adjust our analyses for genetic principal components (to account for possible population admixture). This approach would be ideal, as there may be multiple genetic subpopulations in our samples. Third, while we verified that the NHW and NHB samples aligned with Hardy-Weinberg Equilibrium expectations, the recorded *APOE* genotypes may contain miscalls, which may bias effect estimates, particularly in smaller *APOE* genotype stratified samples. Fourth, a challenge to studying sex differences in AD is that women are more likely than men to survive to older ages.^3^ When a gene, such as *APOE*, has pleiotropic effects on risk for mortality and AD,^49^ this survival bias can cause spurious associations. Finally, given the rarity of *APOE ε2* homozygosity, we were unable to investigate sex differences in allelic dose effects.

In light of the longstanding view that *APOE ε2* protects against AD,^11,27,48,50,51^ our results suggest that in NHW adults *APOE ε2* protects men but not women against cognitive decline. Our findings have important implications for understanding the biological drivers of sex differences in AD risk, which is crucial for developing sex-specific strategies to prevent and treat AD dementia. Large and diverse samples are needed to replicate the present findings and to further clarify the sex-specific effects of *APOE ε2* on risk for AD.

## Data Availability

Data used in the present study (i.e., NACC, ROS/MAP/MARS, ADNI, and Prevent-AD cohorts) is available upon separate requests by investigators to the relevant institutions for each data source.

## 6. Acknowledgements

MEW gratefully acknowledges support from the Canadian Institutes of Health Research (Canada Graduate Scholarships – Master’s). LYX gratefully acknowledges financial support from the Canadian Institutes of Health Research (Doctoral Research Award: Canadian Graduate Scholarships; 202111FBD-476226). RFB is supported by a K99/R00 award from NIA (R00AG061238-03), an Alzheimer’s Association Research Fellowship (AARF-20-675646) and philanthropic support. WS gratefully acknowledges financial support from the Canadian Institute of Health Research (Team Grant); The Natural Sciences and Engineering Research Council of Canada (RGPIN-2017-06962); The Alzheimer’s Association & Brain Canada (AARG501466); Weston Brain Institute & Alzheimer’s Research UK, Alzheimer’s Association and Michael J. Fox Foundation (BAND3). The work was supported in part through funding from the Canada Research Chairs Program. ASPL receives support from R01AG071638, R01AG052488 and RF1AG070436. EN receives support from T32AG000247. RL receives support from K99AG065501. KBC receives support from R01AG072475. CS receives support from the Texas Alzheimer’s Research and Care Consortium (2020-58-81-CR) and R01 AG059727, UF1 NS125513, and P30 AG066546. LLB receives support from R01AG022018 and P30AG010161/P30AG072975. JAS receives support from P30AG010161/P30AG072975, R01AG015819, R01AG17917, R01AG022018. JP receives support from RF1AG054617. JSR receives support from the Harquail Centre for Neuromodulation, the Dr. Sandra Black Centre for Brain Resilience & Recovery, and Canadian Institutes of Health Research (173253, 438475), and the Alzheimer’s Society of Canada.

The NACC database is funded by NIA/NIH Grant U24 AG072122. NACC data are contributed by the NIA-funded ADCs: P50 AG005131 (PI James Brewer, MD, PhD), P50 AG005133 (PI Oscar Lopez, MD), P50 AG005134 (PI Bradley Hyman, MD, PhD), P50 AG005136 (PI Thomas Grabowski, MD), P50 AG005138 (PI Mary Sano, PhD), P50 AG005142 (PI Helena Chui, MD), P50 AG005146 (PI Marilyn Albert, PhD), P50 AG005681 (PI John Morris, MD), P30 AG008017 (PI Jeffrey Kaye, MD), P30 AG008051 (PI Thomas Wisniewski, MD), P50 AG008702 (PI Scott Small, MD), P30 AG010124 (PI John Trojanowski, MD, PhD), P30 AG010129 (PI Charles DeCarli, MD), P30 AG010133 (PI Andrew Saykin, PsyD), P30 AG010161 (PI David Bennett, MD), P30 AG012300 (PI Roger Rosenberg, MD), P30 AG013846 (PI Neil Kowall, MD), P30 AG013854 (PI Robert Vassar, PhD), P50 AG016573 (PI Frank LaFerla, PhD), P50 AG016574 (PI Ronald Petersen, MD, PhD), P30 AG019610 (PI Eric Reiman, MD), P50 AG023501 (PI Bruce Miller, MD), P50 AG025688 (PI Allan Levey, MD, PhD), P30 AG028383 (PI Linda Van Eldik, PhD), P50 AG033514 (PI Sanjay Asthana, MD, FRCP), P30 AG035982 (PI Russell Swerdlow, MD), P50 AG047266 (PI Todd Golde, MD, PhD), P50 AG047270 (PI Stephen Strittmatter, MD, PhD), P50 AG047366 (PI Victor Henderson, MD, MS), P30 AG049638 (PI Suzanne Craft, PhD), P30 AG053760 (PI Henry Paulson, MD, PhD), P30 AG066546 (PI Sudha Seshadri, MD), P20 AG068024 (PI Erik Roberson, MD, PhD), P20 AG068053 (PI Marwan Sabbagh, MD), P20 AG068077 (PI Gary Rosenberg, MD), P20 AG068082 (PI Angela Jefferson, PhD), P30 AG072958 (PI Heather Whitson, MD), P30 AG072959 (PI James Leverenz, MD).

We thank the study participants and staff of the Rush Alzheimer’s Disease Center. This work was supported by NIA grants P30 AG010161 (ROS; PI David Bennett, MD), R01AG17917 (MAP; PI David Bennett, MD) and R01AG22018 (MARS; PI Lisa Barnes, PhD).

