## Supplemental material for "Sex differences in associations between *APOE ε2* and longitudinal cognitive decline"

**Supplementary Material**

**Figure S1. Flow charts depicting sample selection in each cohort.**

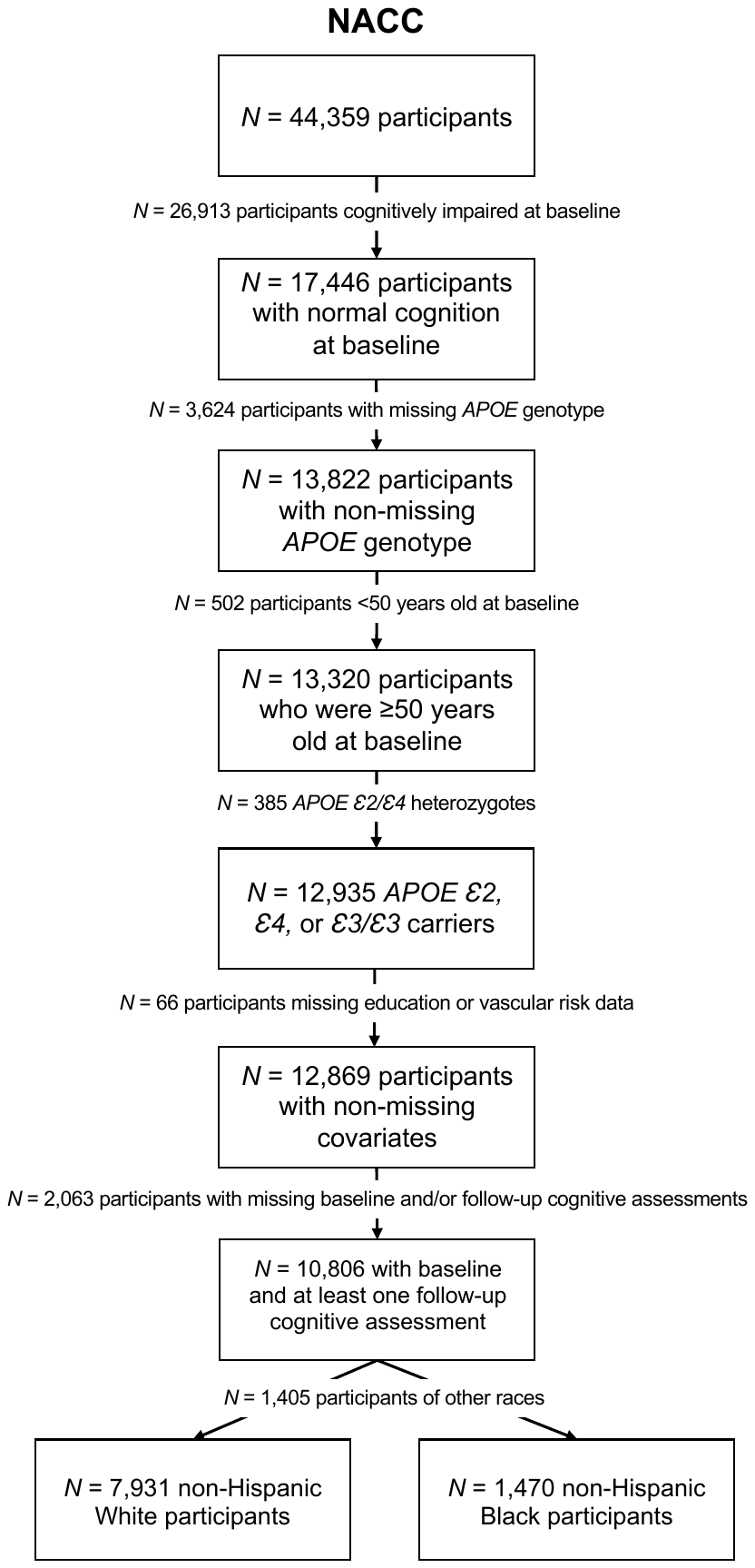

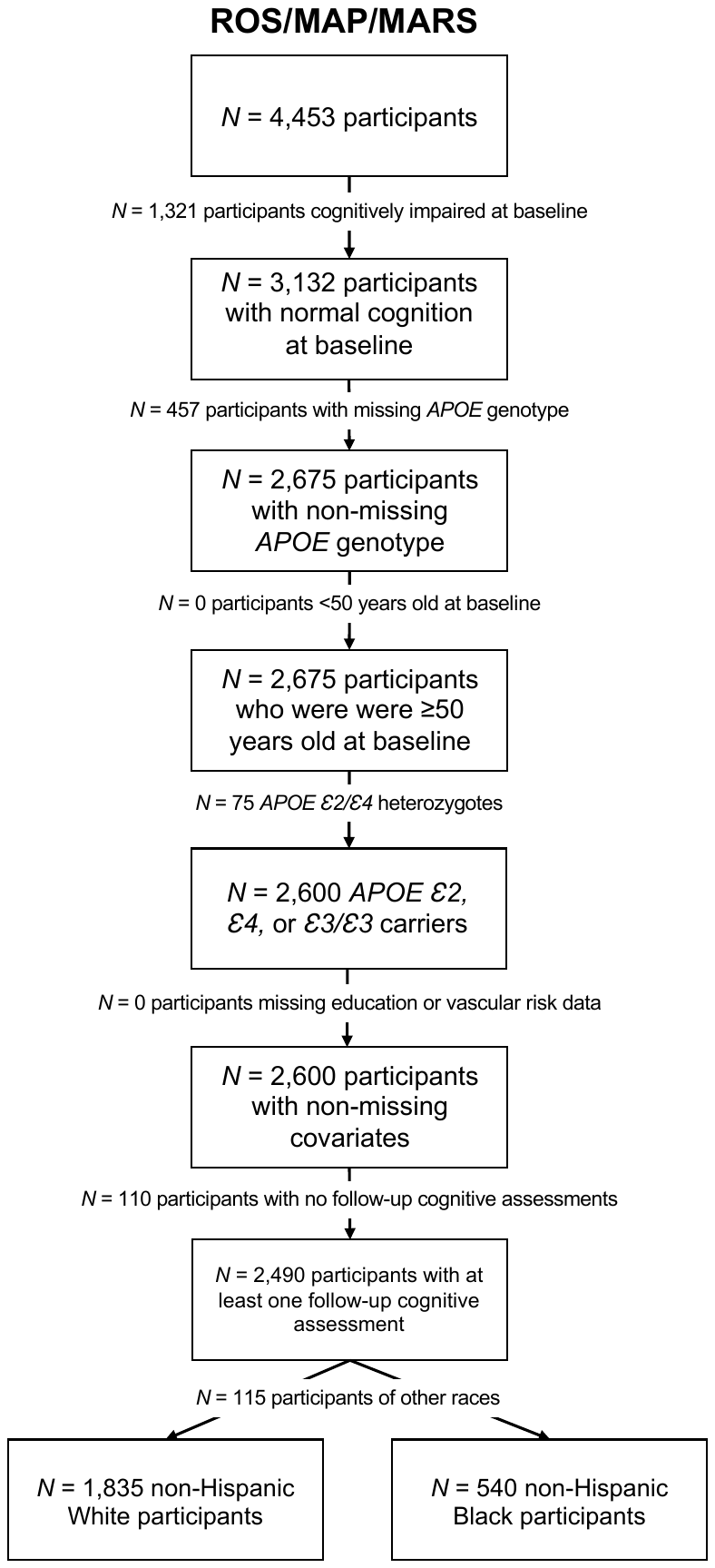

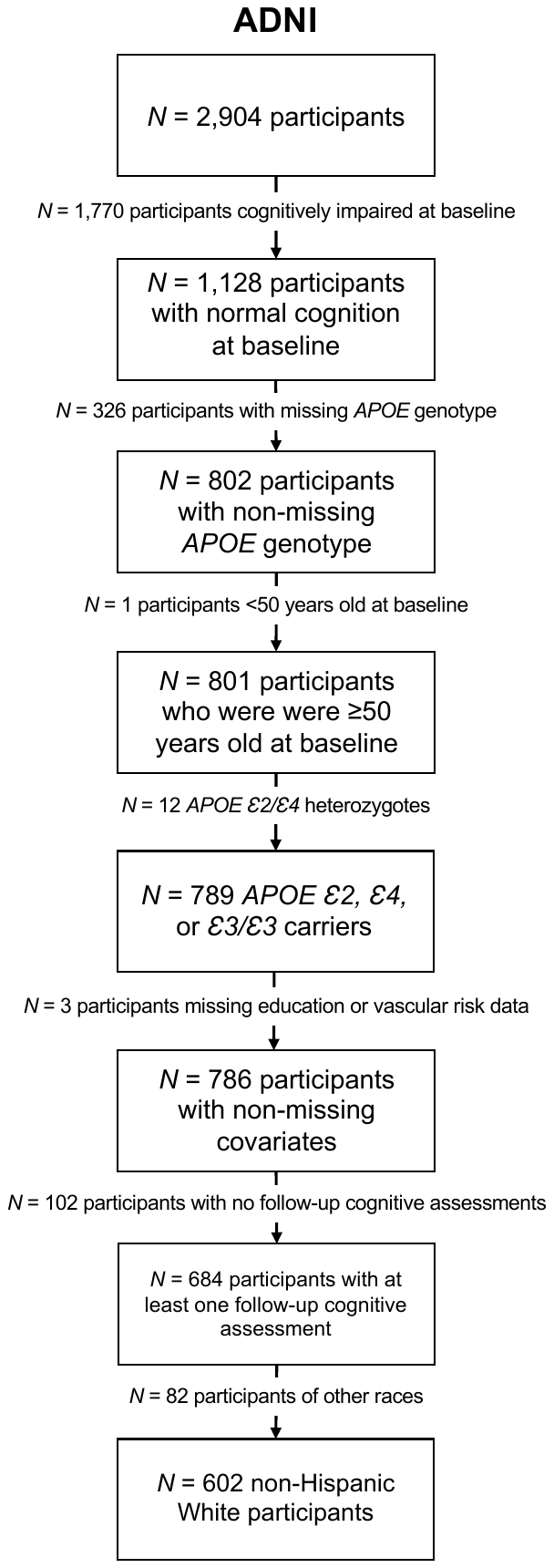

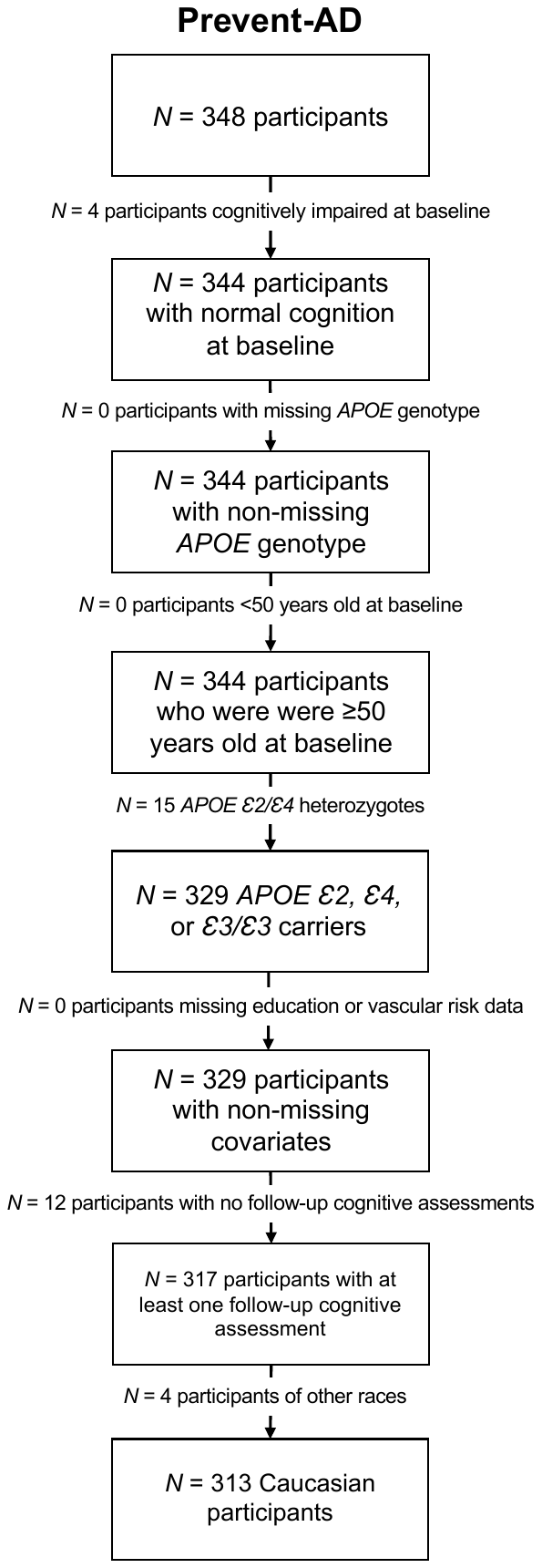

**Additional methods**

**Description of self-reported race and ethnicity for each data source.** In NACC, participants were coded as NHW if they reported their race as “White” and “Hispanic/Latino ethnicity” as “No.” NACC participants were coded as NHB if they reported their race as “Black or African American” and “Hispanic/Latino ethnicity” as “No.” In ROS/MAP/MARS, participants were coded as NHW if they reported their race as “White” and “Spanish/Hispanic/Latino ethnicity” as “No,” and coded as NHB if they reported their race as “Black or African American” and “Spanish/Hispanic/Latino ethnicity” as “No.” In ADNI, participants were coded as NHW if they reported their race as “White” and ethnicity as “Not Hispanic or Latino.” In Prevent-AD, participants were coded as NHW if they reported “Caucasian” ethnicity (no data on Hispanic ethnicity was available).”

**Method for the construction of composite scores of cognition.** In Sample 1 (NACC & ROS/MAP/MARS), we computed a composite score of cognition using tests available in both cohorts: Logical Memory: Immediate and Delayed Recall,^1^ Category Fluency (Animals/Vegetables),^2^ Digit Span^1^and Boston Naming Test.^3^ In 2015, NACC replaced some neuropsychological tests with non-proprietary versions. NACC provides equipercentile equating methods to convert scores on new tests to scores on the older versions,^4^ which we used in the current study.

### In Sample 2, we computed a composite score of cognition using similar tests available in each cohort. In ADNI, cognitive tests included Logical Memory: Delayed Recall,^1^ Rey Auditory Verbal Learning Test: Immediate and Delayed Recall^5^ and Category Fluency (Animals).^2^ In Prevent-AD, cognitive tests included the following tasks from the Repeatable Battery for the Assessment of Neuropsychological Status (RBANS):^6^ Story delayed recall, list delayed recall, list recognition, and semantic fluency.

**Method for the construction of vascular risk scores**. Baseline vascular risk scores were computed as the average of up to 5 dichotomous variables, following a modified procedure previously used in ROS/MAP/MARS.^6^ Included variables were diabetes, hypertension, cholesterol, heart disease, and stroke (depending on data availability in each cohort). If participants had fewer than half of the available items missing, averages were computed; if more than half the items were missing, the score was set to missing (*N* = 2 in NACC; *N* = 0 in ROS/MAP/MARS; *N* = 3 in ADNI; *N* = 0 in Prevent-AD).

In NACC, vascular risk factors were assessed as following (all at baseline): self-reported or clinician-reported histories of hypertension, hypercholesterolemia, diabetes, heart conditions (including heart attack, congestive heart failure, cardiac bypass surgery, and angina), and stroke.

In ROS/MAP/MARS, vascular risk factors were assessed as following (all at baseline): self-reported hypertension or measured systolic blood pressure >= 120 mmHg, LDL cholesterol >= 120 mg/dL, self-reported diabetes or history of taking diabetes medication or measured HBA1c > 6.5 (% of hemoglobin), self-reported or clinician-reported history of heart conditions (including coronary thrombosis, coronary occlusion, myocardial infarction/heart attack, or congestive heart failure) and self-reported or clinician-reported history of stroke.

In ADNI, vascular risk factors were assessed as following (all at baseline): measured systolic blood pressure >= 120 mmHg, total cholesterol >= 200 mg/dL, and self-reported or clinician-reported history of cardiovascular conditions.

In Prevent-AD, vascular risk factors were assessed as following (all at baseline): measured systolic blood pressure >= 120 mmHg or history of treatment for hypertension, LDL cholesterol >= 3.1 mmol/L or history of treatment for hyperlipidemia, measured HBA1c > 6.5 (% of hemoglobin) or history of treatment for diabetes, and history of atrial fibrillation.

**Table S1.** Baseline demographic and clinical characteristics of non-Hispanic Black participants in each cohort.

| **NACC** | | | |
| --- | --- | --- | --- |
| **Variables** | Total sample  (*n* = 1,470) | Women  (*n* = 1,147, 78.0 %) | Men  (*n* = 323, 22.0 %) |
| Age in years, mean (*SD*) | 70.9 (8.07) | 71.1 (8.05) | 70.4 (8.13) |
| Education in years, mean (*SD*) | 14.8 (3.01) | 14.8 (2.93) | 14.9 (3.29) |
| *APOE* *ε2* carriers, *n* (%) | 250 (17.0) | 199 (17.3) | 51 (15.8) |
| *ε2/ε3*, *n* (%) | 233 (15.9) | 187 (16.3) | 46 (14.2) |
| *ε2/ε2*, *n* (%) | 17 (1.12) | 12 (1.05) | 5 (1.55) |
| *APOE* *ε4* carriers, *n* (%) | 506 (34.3) | 376 (32.8)* | 130 (40.2)* |
| *ε3/ε4*, *n* (%) | 458 (31.2) | 341 (29.7) | 117 (36.2) |
| *ε4/ε4*, *n* (%) | 48 (3.27) | 35 (3.05) | 13 (4.02) |
| *APOE ε3/ε3* carriers, *n* (%) | 714 (48.6) | 572 (49.9) | 142 (44.0) |
| Total number of visits, median (*SD*) | 5 (3.05) | 5 (3.08) | 4 (2.93) |
| Progression to AD-related MCI, *n* (%) | 171 (11.6) | 135 (11.8) | 36 (11.1) |
| Progression to AD dementia, *n* (%) | 42 (2.86) | 35 (3.05) | 7 (2.17) |
| **ROS/MAP/MARS** | | | |
| **Variables** | Total sample  (*n* = 540) | Women  (*n* = 436, 80.7%) | Men  (*n* = 134, 19.3%) |
| Age in years, mean (*SD*) | 72.4 (5.98) | 72.3 (6.04) | 72.6 (5.72) |
| Education in years, mean (*SD*) | 15.0 (3.34) | 15.0 (3.24) | 15.0 (3.76) |
| *APOE* *ε2* carriers, *n* (%) | 86 (15.9) | 64 (14.7) | 22 (21.2) |
| *ε2/ε3*, *n* (%) | 83 (15.4) | 61 (14.0) | 22 (21.2) |
| *ε2/ε2*, *n* (%) | 3 (0.56) | 3 (0.69) | 0 (0) |
| *APOE* *ε4* carriers, *n* (%) | 156 (28.9) | 130 (29.8) | 26 (25.0) |
| *ε3/ε4*, *n* (%) | 137 (25.4) | 113 (25.9) | 24 (23.1) |
| *ε4/ε4*, *n* (%) | 19 (3.52) | 17 (3.90) | 2 (1.92) |
| *APOE ε3/ε3* carriers, *n* (%) | 298 (55.2) | 242 (55.5) | 56 (53.8) |
| Total number of visits, median (*SD*) | 8 (4.67) | 8 (4.73) | 8 (4.41) |
| Progression to AD-related MCI, *n* (%) | 133 (27.1) | 102 (25.8) | 31 (32.3) |
| Progression to AD dementia, *n* (%) | 16 (3.26) | 14 (3.54) | 2 (2.08) |

* *p* < .05. P-values represent results of independent samples t-tests and chi-square tests comparing men vs women.

NACC: National Alzheimer’s Coordinating Center cohort

ROS: Religious Orders Study

MAP: Memory Aging Project

MARS: Minority Aging Research Study

SD: standard deviation

MCI: Mild Cognitive Impairment

**Table S2.** Baseline demographic and clinical characteristics of non-Hispanic White participants in each cohort.

| **NACC** | | | |
| --- | --- | --- | --- |
| **Variables** | Total sample  (*n* = 7,931) | Women  (*n* = 4,980, 62.8 %) | Men  (*n* = 2,851, 37.2%) |
| Age in years, mean (*SD*) | 71.9 (9.08) | 71.7 (9.17)* | 72.4 (8.91)* |
| Education in years, mean (*SD*) | 16.2 (2.63) | 15.9 (2.54)* | 16.8 (2.69)* |
| *APOE* *ε2* carriers, *n* (%) | 995 (12.5) | 645 (13.0) | 350 (11.9) |
| *ε2/ε3*, *n* (%) | 956 (12.1) | 626 (12.6) | 330 (11.2) |
| *ε2/ε2*, *n* (%) | 39 (0.49) | 19 (0.38) | 20 (0.68) |
| *APOE* *ε4* carriers, *n* (%) | 2249 (28.4) | 1391(27.9) | 858 (29.1) |
| *ε3/ε4*, *n* (%) | 2012 (25.4) | 1249 (25.1) | 763 (25.9) |
| *ε4/ε4*, n (%) | 237 (2.99) | 142 (2.85) | 95 (3.22) |
| *APOE ε3/ε3* carriers, *n* (%) | 4687 (59.1) | 2944 (59.1) | 1743 (59.1) |
| Total number of visits, median (*SD*) | 5 (3.40) | 5 (3.42) | 5 (3.35) |
| Progression to AD-related MCI, *n* (%) | 1214 (15.3) | 740 (14.9) | 474 (16.1) |
| Progression to AD dementia, *n* (%) | 247 (3.11) | 154 (3.09) | 93 (3.15) |
| **ROS/MAP** | | | |
| **Variables** | Total sample  (*n* = 1,835) | Women  (*n* = 1,364, 74.3%) | Men  (*n* = 471, 25.7%) |
| Age in years, mean (*SD*) | 77.3 (7.20) | 77.7 (7.30)* | 76.2 (6.80)* |
| Education in years, mean (*SD*) | 16.6 (3.57) | 16.3 (3.38)* | 17.5 (3.92)* |
| *APOE* *ε2* carriers, *n* (%) | 265 (14.4) | 195 (14.3) | 70 (14.9) |
| *ε2/ε3*, *n* (%) | 255 (13.9) | 188 (13.8) | 67 (14.2) |
| *ε2/ε2*, *n* (%) | 10 (0.54) | 7 (0.51) | 3 (0.64) |
| *APOE* *ε4* carriers, *n* (%) | 373 (20.3) | 279 (20.5) | 94 (20.0) |
| *ε3/ε4*, *n* (%) | 350 (19.1) | 259 (19.0) | 91 (19.3) |
| *ε4/ε4*, n (%) | 23 (1.25) | 20 (1.47) | 3 (0.64) |
| *APOE ε3/ε3* carriers, *n* (%) | 1197 (65.2) | 890 (65.2) | 307 (65.1) |
| Total number of visits, median (*SD*) | 10 (5.79) | 10 (5.63) | 10 (6.07) |
| Progression to AD-related MCI, *n* (%) | 557 (31.6) | 398 (30.4) | 159 (35.3) |
| Progression to AD dementia, *n* (%) | 183 (10.4) | 137 (10.5) | 46 (10.2) |
| **ADNI** | | | |
| **Variables** | Total sample  (*n* = 602) | Women  (*n* = 318, 52.8 %) | Men  (*n* = 284, 47.2%) |
| Age in years, mean (*SD*) | 73.5 (5.96) | 72.6 (5.92)* | 74.4 (5.86)* |
| Education in years, mean (*SD*) | 16.6 (2.55) | 16.1 (2.57)* | 17.1 (2.39)* |
| *APOE* *ε2* carriers, *n* (%) | 70 (11.6) | 32 (10.1) | 38 (13.3) |
| *ε2/ε3*, *n* (%) | 69 (11.5) | 32 (10.1) | 37 (13.0) |
| *ε2/ε2*, *n* (%) | 1 (0.17) | 0 (0) | 1 (0.35) |
| *APOE* *ε4* carriers, *n* (%) | 174 (28.9) | 99 (31.1) | 75 (26.3) |
| *ε3/ε4*, *n* (%) | 157 (26.1) | 89 (28.0) | 68 (23.9) |
| *ε4/ε4*, n (%) | 17 (2.82) | 10 (3.15) | 7 (2.46) |
| *APOE ε3/ε3* carriers, *n* (%) | 358 (59.5) | 187 (58.8) | 172 (60.4) |
| Total number of visits, median (*SD*) | 5 (3.17) | 5 (3.03)* | 5 (3.30)* |
| Progression to MCI, *n* (%) | 106 (17.6) | 50 (15.7) | 56 (19.7) |
| Progression to AD dementia, *n* (%) | 5 (0.83) | 2 (0.63) | 3 (1.06) |
| **Prevent-AD** | | | |
| **Variables** | Total sample  (*n* = 313) | Women  (*n* = 224, 71.6 %) | Men  (*n* = 89, 28.4%) |
| Age in years, mean (*SD*) | 63.5 (5.02) | 63.4 (5.01) | 63.8 (5.03) |
| Education in years, mean (*SD*) | 15.4 (3.40) | 15.2 (3.37) | 16.0 (3.43) |
| *APOE* *ε2* carriers, *n* (%) | 38 (12.1) | 23 (10.3) | 15 (16.9) |
| *ε2/ε3*, *n* (%) | 38 (12.1) | 23 (10.3) | 15 (16.9) |
| *ε2/ε2*, *n* (%) | 0 (0) | 0 (0) | 0 (0) |
| *APOE* *ε4* carriers, *n* (%) | 113 (36.1) | 77 (34.4) | 36 (40.4) |
| *ε3/ε4*, *n* (%) | 106 (33.9) | 71 (31.7) | 35 (39.3) |
| *ε4/ε4*, n (%) | 7 (2.24) | 6 (26.8) | 1 (1.12) |
| *APOE ε3/ε3* carriers, *n* (%) | 162 (51.8) | 124 (55.4) | 38 (42.7) |
| Total number of visits, median (*SD*) | 6 (2.13) | 6 (2.11) | 6 (2.18) |
| Progression to probable MCI, *n* (%) | 37 (11.8) | 24 (10.7) | 13 (14.6) |

* *p* < .05. P-values represent results of independent samples t-tests and chi-square tests comparing men vs women.

NACC: National Alzheimer’s Coordinating Center cohort

ROS: Religious Orders Study

MAP: Memory Aging Project

MARS: Minority Aging Research Study

ADNI: Alzheimer’s Disease Neuroimaging Initiative

SD: standard deviation

MCI: Mild Cognitive Impairment

**Table S3. Full regression outputs of linear mixed models examining sex differences in associations between *APOE Ɛ2* (vs. *Ɛ3/Ɛ3*) and longitudinal cognition in non-Hispanic Black participants in Sample 1 (NACC & ROS/MAP/MARS).**

|  | | Standardized *β* estimate | | 95% confidence interval | | *P* value |
| --- | --- | --- | --- | --- | --- | --- |
| **Cognition ~ sex × *APOE* × time + [baseline age + education + baseline vascular risk score + cohort] × time +** $\boldsymbol{\surd}$**visit number + time^2^; *N* = 2,010** | | | | | | |
| Sex (male) × *APOE* (*Ɛ2*) × time | | -0.011 | | -0.153 – 0.131 | | .88 |
| Sex (male) × *APOE* (*Ɛ4*) × time | | 0.103 | | -0.017 – 0.223 | | .09 |
| Sex (male) × time | | -0.019 | | -0.096 – 0.058 | | .63 |
| *APOE* (*Ɛ2*) × time | | 0.050 | | -0.015 – 0.115 | | .13 |
| *APOE* (*Ɛ4*) × time | | -0.075 | | -0.129 – -0.021 | | .006 |
| Sex (male) × *APOE* (*Ɛ2*) | | 0.056 | | -0.188 – 0.301 | | .65 |
| Sex (male) × *APOE* (*Ɛ4*) | | 0.046 | | -0.149 – 0.241 | | .64 |
| Baseline age × time | | -0.103 | | -0.124 – -0.081 | | <.001 |
| Education × time | | -0.005 | | -0.027 – 0.017 | | .65 |
| Vascular risk × time | | -0.028 | | -0.050 – -0.006 | | .01 |
| Cohort (ROS/MAP/MARS) × time | | 0.054 | | 0.006 – 0.103 | | .03 |
| Time^2^ | | -0.070 | | -0.081 – -0.059 | | <.001 |
| Sex (male) | | -0.093 | | -0.221 – 0.034 | | .15 |
| *APOE* (*Ɛ2*) | | 0.003 | | -0.109 – 0.116 | | .96 |
| *APOE* (*Ɛ4*) | | -0.083 | | -0.174 – 0.007 | | .07 |
| Baseline age | | -0.356 | | -0.391 – -0.322 | | <.001 |
| Education | | 0.329 | | 0.292 – 0.366 | | <.001 |
| $\surd$visit number | | 0.106 | | 0.066 – 0.146 | | <.001 |
| Vascular risk | | -0.058 | | -0.095 – -0.021 | | .002 |
| Cohort (ROS/MAP/MARS) | | 0.134 | | 0.051 – 0.218 | | .002 |
| Time | | -0.190 | | -0.252 – -0.127 | | <.001 |
| **Cognition ~ *APOE* × time + [baseline age + education + baseline vascular risk score + cohort] × time +** $\boldsymbol{\surd}$**visit number + time^2^; Men; *N* = 427** | | | | | | |
| *APOE* (*Ɛ2*) × time | 0.037 | | -0.077 – 0.151 | | .52 | |
| *APOE* (*Ɛ4*) × time | 0.012 | | -0.087 – 0.110 | | .81 | |
| Baseline age × time | -0.078 | | -0.122 – -0.034 | | <.001 | |
| Education × time | -0.028 | | -0.072 – 0.017 | | .22 | |
| Vascular risk × time | -0.020 | | -0.067 – 0.027 | | .41 | |
| Cohort (ROS/MAP/MARS) × time | -0.025 | | -0.129 – 0.079 | | .64 | |
| Time^2^ | -0.022 | | -0.048 – 0.003 | | .09 | |
| *APOE* (*Ɛ2*) | 0.057 | | -0.165 – 0.280 | | .61 | |
| *APOE* (*Ɛ4*) | -0.056 | | -0.236 – 0.123 | | .54 | |
| Baseline age | -0.353 | | -0.430 – -0.277 | | <.001 | |
| Education | 0.334 | | 0.253 – 0.415 | | <.001 | |
| $\surd$visit number | 0.158 | | 0.067 – 0.248 | | .001 | |
| Vascular risk | -0.053 | | -0.136 – 0.031 | | .22 | |
| Cohort (ROS/MAP/MARS) | 0.046 | | -0.150 – 0.242 | | .65 | |
| Time | -0.211 | | -0.348 – -0.074 | | .003 | |
| **Cognition ~ *APOE* × time + [baseline age + education + baseline vascular risk score + cohort] × time +** $\boldsymbol{\surd}$**visit number + time^2^; Women; *N* = 1,583** | | | | | | |
| *APOE* (*Ɛ2*) × time | 0.053 | | -0.014 – 0.119 | | .12 | |
| *APOE* (*Ɛ4*) × time | -0.076 | | -0.131 – -0.021 | | .007 | |
| Baseline age × time | -0.107 | | -0.132 – -0.083 | | <.001 | |
| Education × time | 0.001 | | -0.024 – 0.025 | | .95 | |
| Vascular risk × time | -0.029 | | -0.054 – -0.004 | | .03 | |
| Cohort (ROS/MAP) × time | 0.071 | | 0.017 – 0.126 | | .01 | |
| Time^2^ | -0.079 | | -0.091 – -0.067 | | <.001 | |
| *APOE* (*Ɛ2*) | 0.006 | | -0.106 – 0.118 | | .92 | |
| *APOE* (*Ɛ4*) | -0.084 | | -0.174 – 0.006 | | .07 | |
| Baseline age | -0.356 | | -0.394 – -0.318 | | <.001 | |
| Education | 0.328 | | 0.287 – 0.370 | | <.001 | |
| $\surd$visit number | 0.098 | | 0.053 – 0.143 | | <.001 | |
| Vascular risk | -0.058 | | -0.100 – -0.017 | | .006 | |
| Cohort (ROS/MAP/MARS) | 0.152 | | 0.059 – 0.245 | | .001 | |
| Time | -0.193 | | -0.262 – -0.125 | | <.001 | |
| **Cognition ~ sex × time + [baseline age + education + baseline vascular risk score + cohort] × time +** $\boldsymbol{\surd}$**visit number + time^2^; *APOE* *Ɛ4*** **carriers; *N* = 662** | | | | | | |
| Sex (male) × time | 0.081 | | -0.008 – 0.169 | | .07 | |
| Baseline age × time | -0.091 | | -0.128 – -0.054 | | <.001 | |
| Education × time | 0.003 | | -0.035 – 0.041 | | .86 | |
| Vascular risk × time | -0.031 | | -0.069 – 0.007 | | .11 | |
| Cohort (ROS/MAP/MARS) × time | 0.055 | | -0.028 – 0.139 | | .19 | |
| Time^2^ | -0.080 | | -0.100 – -0.061 | | <.001 | |
| Sex (male) | -0.055 | | -0.200 – 0.091 | | .46 | |
| Baseline age | -0.359 | | -0.418 – -0.299 | | <.001 | |
| Education | 0.366 | | 0.302 – 0.430 | | <.001 | |
| $\surd$visit number | 0.060 | | -0.010 – 0.130 | | .09 | |
| Vascular risk | -0.012 | | -0.074 – 0.049 | | .70 | |
| Cohort (ROS/MAP/MARS) | 0.120 | | -0.027 – 0.268 | | .11 | |
| Time | -0.181 | | -0.281 – -0.082 | | <.001 | |
| **Cognition ~ *APOE* × time + [sex + baseline age + education + baseline vascular risk score + cohort] × time +** $\boldsymbol{\surd}$**visit number + time^2^; *N* = 2,010** | | | | | | |
| *APOE* (*Ɛ2*) × time | 0.046 | | -0.012 – 0.104 | | .12 | |
| *APOE* (*Ɛ4*) × time | -0.054 | | -0.102 – -0.006 | | .03 | |
| Sex (male) × time | 0.013 | | -0.040 – 0.066 | | .63 | |
| Baseline age × time | -0.103 | | -0.124 – -0.082 | | <.001 | |
| Education × time | -0.006 | | -0.028 – 0.015 | | .58 | |
| Vascular risk × time | -0.028 | | -0.050 – -0.005 | | .02 | |
| Cohort (ROS/MAP/MARS) × time | 0.052 | | 0.004 – 0.100 | | .03 | |
| Time^2^ | -0.070 | | -0.081 – -0.058 | | <.001 | |
| *APOE* (*Ɛ2*) | 0.014 | | -0.086 – 0.114 | | .78 | |
| *APOE* (*Ɛ4*) | -0.075 | | -0.155 – 0.006 | | .07 | |
| Sex (male) | -0.069 | | -0.157 – 0.018 | | .12 | |
| Baseline age | -0.357 | | -0.391 – -0.323 | | <.001 | |
| Education | 0.329 | | 0.292 – 0.365 | | <.001 | |
| $\surd$visit number | 0.106 | | 0.066 – 0.146 | | <.001 | |
| Vascular risk | -0.058 | | -0.095 – -0.021 | | .002 | |
| Cohort (ROS/MAP/MARS) | 0.134 | | 0.051 – 0.218 | | .002 | |
| Time | -0.194 | | -0.256 – -0.133 | | <.001 | |

**Table S4. Full regression outputs of linear mixed models examining sex differences in associations between *APOE Ɛ2* (vs. *Ɛ3/Ɛ3*) and longitudinal cognition in non-Hispanic White participants in Sample 1 (NACC & ROS/MAP).**

|  | Standardized *β* estimate | 95% confidence interval | *P* value |
| --- | --- | --- | --- |
| **Cognition ~ sex × *APOE* × time + [baseline age + education + baseline vascular risk score + cohort] × time +** $\boldsymbol{\surd}$**visit number + time^2^; *N* = 9,766** | | | |
| Sex (male) × *APOE* (*Ɛ2*) × time | 0.097 | 0.023 – 0.172 | .01 |
| Sex (male) × *APOE* (*Ɛ4*) × time | 0.064 | 0.007 – 0.120 | .03 |
| Sex (male) × time | 0.000 | -0.032 – 0.032 | >.99 |
| *APOE* (*Ɛ2*) × time | -0.001 | -0.044 – 0.041 | .95 |
| *APOE* (*Ɛ4*) × time | -0.192 | -0.226 – -0.159 | <.001 |
| Sex (male) × *APOE* (*Ɛ2*) | 0.130 | 0.020 – 0.240 | .02 |
| Sex (male) × *APOE* (*Ɛ4*) | 0.047 | -0.036 – 0.129 | .27 |
| Baseline age × time | -0.205 | -0.217 – -0.193 | <.001 |
| Education × time | 0.006 | -0.006 – 0.018 | .31 |
| Vascular risk × time | -0.002 | -0.015 – 0.011 | .78 |
| Cohort (ROS/MAP) × time | 0.047 | 0.017 – 0.078 | .002 |
| Time^2^ | -0.132 | -0.137 – -0.127 | <.001 |
| Sex (male) | -0.191 | -0.237 – -0.144 | <.001 |
| *APOE* (*Ɛ2*) | -0.002 | -0.066 – 0.061 | .95 |
| *APOE* (*Ɛ4*) | -0.239 | -0.288 – -0.190 | <.001 |
| Baseline age | -0.426 | -0.443 – -0.409 | <.001 |
| Education | 0.225 | 0.208 – 0.243 | <.001 |
| $\surd$visit number | 0.102 | 0.085 – 0.120 | <.001 |
| Vascular risk | -0.032 | -0.050 – -0.013 | .001 |
| Cohort (ROS/MAP) | -0.138 | -0.185 – -0.092 | <.001 |
| Time | -0.291 | -0.321 – -0.261 | <.001 |
| **Cognition ~ *APOE* × time + [baseline age + education + baseline vascular risk score + cohort] × time +** $\boldsymbol{\surd}$**visit number + time^2^; Men; *N* = 3,422** | | | |
| *APOE* (*Ɛ2*) × time | 0.096 | 0.037 – 0.155 | .001 |
| *APOE* (*Ɛ4*) × time | -0.127 | -0.171 – -0.083 | <.001 |
| Baseline age × time | -0.189 | -0.208 – -0.170 | <.001 |
| Education × time | -0.005 | -0.025 – 0.014 | .60 |
| Vascular risk × time | -0.002 | -0.023 – 0.018 | .82 |
| Cohort (ROS/MAP) × time | 0.052 | -0.000 – 0.105 | .05 |
| Time^2^ | -0.099 | -0.108 – -091 | <.001 |
| *APOE* (*Ɛ2*) | 0.132 | 0.041 – 0.224 | .005 |
| *APOE* (*Ɛ4*) | -0.197 | -0.265 – -0.129 | <.001 |
| Baseline age | -0.419 | -0.448 – -0.391 | <.001 |
| Education | 0.246 | 0.216 – 0.277 | <.001 |
| $\surd$visit number | 0.149 | 0.119 – 0.179 | <.001 |
| Vascular risk | -0.019 | -0.050 – 0.013 | .24 |
| Cohort (ROS/MAP) | -0.203 | -0.291 – -0.116 | <.001 |
| Time | -0.336 | -0.382 – -290 | <.001 |
| **Cognition ~ *APOE* × time + [baseline age + education + baseline vascular risk score + cohort] × time +** $\boldsymbol{\surd}$**visit number + time^2^; Women; *N* = 6,344** | | | |
| *APOE* (*Ɛ2*) × time | -0.001 | -0.044 – 0.043 | .97 |
| *APOE* (*Ɛ4*) × time | -0.192 | -0.227 – -0.158 | <.001 |
| Baseline age × time | -0.212 | -0.227 – -0.197 | <.001 |
| Education × time | 0.011 | -0.004 – 0.026 | .15 |
| Vascular risk × time | -0.001 | -0.018 – 0.016 | .90 |
| Cohort (ROS/MAP) × time | 0.045 | 0.008 – 0.083 | .02 |
| Time^2^ | -0.147 | -0.153 – -0.141 | <.001 |
| *APOE* (*Ɛ2*) | -0.003 | -0.066 – 0.060 | .92 |
| *APOE* (*Ɛ4*) | -0.238 | -0.287 – -0.188 | <.001 |
| Baseline age | -0.430 | -0.452 – -0.408 | <.001 |
| Education | 0.211 | 0.190 – 0.233 | <.001 |
| $\surd$visit number | 0.080 | 0.058 – 0.102 | <.001 |
| Vascular risk | -0.040 | -0.063 – -0.016 | .001 |
| Cohort (ROS/MAP) | -0.107 | -0.163 – -0.052 | <.001 |
| Time | -0.270 | -0.305 – -0.235 | <.001 |
| **Cognition ~ sex × time + [baseline age + education + baseline vascular risk score + cohort] × time +** $\boldsymbol{\surd}$**visit number + time^2^; *APOE* *Ɛ2*** **carriers; *N* = 1,260** | | | |
| Sex (male) × time | 0.120 | 0.051 – 0.190 | **.001** |
| Baseline age × time | -0.189 | -0.221 – -0.156 | **<.001** |
| Education × time | -0.017 | -0.050 – 0.015 | .30 |
| Vascular risk × time | -0.046 | -0.083 – -0.008 | **.02** |
| Cohort (ROS/MAP) × time | 0.116 | 0.032 – 0.201 | **.007** |
| Time^2^ | -0.110 | -0.124 – -0.096 | **<.001** |
| Sex (male) | -0.047 | -0.150 – 0.055 | .36 |
| Baseline age | -0.407 | -0.454 – -0.360 | **<.001** |
| Education | 0.207 | 0.158 – 0.256 | **<.001** |
| $\surd$visit number | 0.139 | 0.088 – 0.190 | **<.001** |
| Vascular risk | -0.070 | -0.125 – -0.015 | **.01** |
| Cohort (ROS/MAP) | -0.089 | -0.218 – 0.040 | .17 |
| Time | -0.408 | -0.487 – -0.329 | **<.001** |
| **Cognition ~ sex × time + [baseline age + education + baseline vascular risk score + cohort] × time +** $\boldsymbol{\surd}$**visit number + time^2^; *APOE* *Ɛ3/Ɛ3*** **carriers; *N* = 5,884** | | | |
| Sex (male) × time | -0.000 | -0.031 – 0.030 | .99 |
| Baseline age × time | -0.207 | -0.222 – -0.193 | **<.001** |
| Education × time | 0.012 | -0.003 – 0.026 | .13 |
| Vascular risk × time | 0.008 | -0.008 – 0.024 | .32 |
| Cohort (ROS/MAP) × time | 0.052 | 0.015 – 0.088 | **.005** |
| Time^2^ | -0.137 | -0.143 – -0.131 | **<.001** |
| Sex (male) | -0.195 | -0.241 – -0.149 | **<.001** |
| Baseline age | -0.425 | -0.447 – -0.404 | **<.001** |
| Education | 0.238 | 0.215 – 0.260 | **<.001** |
| $\surd$visit number | 0.090 | 0.066 – 0.113 | **<.001** |
| Vascular risk | -0.032 | -0.056 – -0.009 | **.007** |
| Cohort (ROS/MAP) | -0.141 | -0.198 – -0.084 | **<.001** |
| Time | -0.299 | -0.334 – -0.263 | **<.001** |
| **Cognition ~ sex × time + [baseline age + education + baseline vascular risk score + cohort] × time +** $\boldsymbol{\surd}$**visit number + time^2^; *APOE* *Ɛ4*** **carriers; *N* = 2,622** | | | |
| Sex (male) × time | 0.053 | 0.002 – 0.104 | .04 |
| Baseline age × time | -0.196 | -0.220 – -0.171 | <.001 |
| Education × time | 0.007 | -0.017 – 0.032 | .56 |
| Vascular risk × time | -0.007 | -0.033 – 0.019 | .59 |
| Cohort (ROS/MAP) × time | 0.001 | -0.068 – 0.070 | .98 |
| Time^2^ | -0.134 | -0.143 – -0.125 | <.001 |
| Sex (male) | -0.141 | -0.213 – -0.069 | <.001 |
| Baseline age | -0.419 | -0.453 – -0.385 | <.001 |
| Education | 0.208 | 0.173 – 0.244 | <.001 |
| $\surd$visit number | 0.103 | 0.071 – 0.135 | <.001 |
| Vascular risk | -0.017 | -0.054 – 0.020 | .36 |
| Cohort (ROS/MAP) | -0.144 | -0.246 – -0.042 | .006 |
| Time | -0.387 | -0.440 – -0.335 | **<.001** |

**Table S5. Full regression outputs of linear mixed models examining sex differences in associations between *APOE Ɛ2* (vs. *Ɛ3/Ɛ3*) and longitudinal cognition in non-Hispanic White NACC participants.**

|  | Standardized *β* estimate | 95% confidence interval | *P* value |
| --- | --- | --- | --- |
| **Cognition ~ sex × *APOE* × time + [baseline age + education + baseline vascular risk score] × time +** $\boldsymbol{\surd}$**visit number + time^2^; *N* = 7,931** | | | |
| Sex (male) × *APOE* (*Ɛ2*) × time | 0.081 | 0.010 – 0.152 | .02 |
| Sex (male) × *APOE* (*Ɛ4*) × time | 0.042 | -0.009 – 0.094 | .11 |
| Sex (male) × time | -0.002 | -0.032 – 0.028 | .88 |
| *APOE* (*Ɛ2*) × time | -0.008 | -0.050 – 0.034 | .70 |
| *APOE* (*Ɛ4*) × time | -0.133 | -0.165 – -0.101 | <.001 |
| Sex (male) × *APOE* (*Ɛ2*) | 0.154 | 0.032 – 0.277 | .01 |
| Sex (male) × *APOE* (*Ɛ4*) | 0.055 | -0.033 – 0.144 | .22 |
| Baseline age × time | -0.164 | -0.175 – -0.153 | <.001 |
| Education × time | 0.007 | -0.004 – 0.018 | .22 |
| Vascular risk × time | 0.002 | -0.010 – 0.013 | .77 |
| Time^2^ | -0.093 | -0.100 – -0.087 | <.001 |
| Sex (male) | -0.229 | -0.280 – -0.177 | <.001 |
| *APOE* (*Ɛ2*) | -0.025 | -0.098 – 0.048 | .50 |
| *APOE* (*Ɛ4*) | -0.215 | -0.270 – -0.160 | <.001 |
| Baseline age | -0.419 | -0.438 – -0.400 | <.001 |
| Education | 0.251 | 0.231 – 0.270 | <.001 |
| $\surd$visit number | 0.091 | 0.071 – 0.111 | <.001 |
| Vascular risk | -0.029 | -0.048 – -0.010 | .003 |
| Time | -0.187 | -0.216 – -0.147 | <.001 |
| **Cognition ~ *APOE* × time + [baseline age + education + baseline vascular risk score] × time +** $\boldsymbol{\surd}$**visit number + time^2^; Men; *N* = 2,951** | | | |
| *APOE* (*Ɛ2*) × time | 0.074 | 0.020 – 0.128 | .008 |
| *APOE* (*Ɛ4*) × time | -0.090 | -0.129 – -0.051 | <.001 |
| Baseline age × time | -0.155 | -0.172 – -0.138 | <.001 |
| Education × time | 0.004 | -0.014 – 0.021 | .68 |
| Vascular risk × time | 0.005 | -0.013 – 0.022 | .59 |
| Time^2^ | -0.078 | -0.089 – -0.067 | <.001 |
| *APOE* (*Ɛ2*) | 0.134 | 0.035 – 0.232 | .008 |
| *APOE* (*Ɛ4*) | -0.162 | -0.233 – -0.091 | <.001 |
| Baseline age | -0.415 | -0.445 – -0.384 | <.001 |
| Education | 0.254 | 0.223 – 0.284 | <.001 |
| $\surd$visit number | 0.109 | 0.075 – 0.142 | <.001 |
| Vascular risk | -0.006 | -0.037 – 0.025 | .71 |
| Time | -0.210 | -0.255 – -0.165 | <.001 |
| **Cognition ~ *APOE* × time + [baseline age + education + baseline vascular risk score] × time +** $\boldsymbol{\surd}$**visit number + time^2^; Women; *N* = 4,980** | | | |
| *APOE* (*Ɛ2*) × time | -0.008 | -0.051 – 0.035 | .71 |
| *APOE* (*Ɛ4*) × time | -0.135 | -0.168 – -0.102 | **<.001** |
| Baseline age × time | -0.169 | -0.184 – -0.154 | **<.001** |
| Education × time | 0.009 | -0.006 – 0.023 | .23 |
| Vascular risk × time | 0.000 | -0.015 – 0.015 | .99 |
| Time^2^ | -0.102 | -0.111 – -0.094 | **<.001** |
| *APOE* (*Ɛ2*) | -0.029 | -0.103 – 0.044 | .44 |
| *APOE* (*Ɛ4*) | -0.216 | -0.272 – -0.161 | **<.001** |
| Baseline age | -0.422 | -0.446 – -0.397 | **<.001** |
| Education | 0.244 | 0.219 – 0.268 | **<.001** |
| $\surd$visit number | 0.082 | 0.056 – 0.107 | **<.001** |
| Vascular risk | -0.044 | -0.069 – -0.020 | **<.001** |
| Time | -0.175 | -0.210 – -0.139 | **<.001** |
| **Cognition ~ sex × time + [baseline age + education + baseline vascular risk score] × time +** $\boldsymbol{\surd}$**visit number + time^2^; *APOE* *Ɛ2*** **carriers; *N* = 995** | | | |
| Sex (male) × time | 0.095 | 0.028 – 0.161 | .005 |
| Baseline age × time | -0.147 | -0.178 – -0.115 | <.001 |
| Education × time | -0.016 | -0.048 – 0.015 | .31 |
| Vascular risk × time | -0.024 | -0.056 – 0.008 | .15 |
| Time^2^ | -0.094 | -0.114 – -0.075 | <.001 |
| Sex (male) | -0.061 | -0.175 – 0.054 | .30 |
| Baseline age | -0.408 | -0.461 – -0.356 | <.001 |
| Education | 0.214 | 0.160 – 0.267 | <.001 |
| $\surd$visit number | 0.081 | 0.023 – 0.139 | .007 |
| Vascular risk | -0.058 | -0.112 – -0.003 | .04 |
| Time | -0.221 | -0.299 – -0.142 | <.001 |
| **Cognition ~ sex × time + [baseline age + education + baseline vascular risk score] × time +** $\boldsymbol{\surd}$**visit number + time^2^; *APOE* *Ɛ3/Ɛ3*** **carriers; *N* = 4,687** | | | |
| Sex (male) × time | -0.005 | -0.033 – 0.024 | .75 |
| Baseline age × time | -0.163 | -0.177 – -0.149 | <.001 |
| Education × time | 0.014 | 0.001 – 0.028 | .04 |
| Vascular risk × time | 0.005 | -0.008 – 0.019 | .44 |
| Time^2^ | -0.090 | -0.099 – -0.081 | <.001 |
| Sex (male) | -0.234 | -0.285 – -0.183 | <.001 |
| Baseline age | -0.407 | -0.431 – -0.383 | <.001 |
| Education | 0.260 | 0.235 – 0.284 | <.001 |
| $\surd$visit number | 0.090 | 0.064 – 0.117 | <.001 |
| Vascular risk | -0.038 | -0.062 – -0.013 | .003 |
| Time | -0.197 | -0.233 – -0.162 | <.001 |

**Table S6. Full regression outputs of linear mixed models examining sex differences in associations between *APOE Ɛ2* (vs. *Ɛ3/Ɛ3*) and longitudinal cognition in non-Hispanic White ROS/MAP participants.**

|  | Standardized *β* estimate | 95% confidence interval | *P* value |
| --- | --- | --- | --- |
| **Cognition ~ sex × *APOE* × time + [baseline age + education + baseline vascular risk score] × time +** $\boldsymbol{\surd}$**visit number + time^2^; *N* = 1,835** | | | |
| Sex (male) × *APOE* (*Ɛ2*) × time | 0.127 | -0.069 – 0.323 | .20 |
| Sex (male) × *APOE* (*Ɛ4*) × time | 0.007 | -0.171 – 0.185 | .94 |
| Sex (male) × time | 0.039 | -0.048 – 0.126 | .37 |
| *APOE* (*Ɛ2*) × time | 0.014 | -0.089 – 0.117 | .79 |
| *APOE* (*Ɛ4*) × time | -0.293 | -0.382 – -0.204 | <.001 |
| Sex (male) × *APOE* (*Ɛ2*) | 0.085 | -0.190 – 0.359 | .55 |
| Sex (male) × *APOE* (*Ɛ4*) | -0.099 | -0.346 – 0.147 | .43 |
| Baseline age × time | -0.256 | -0.288 – -0.224 | <.001 |
| Education × time | -0.021 | -0.052 – 0.011 | .20 |
| Vascular risk × time | -0.021 | -0.051 – 0.010 | .18 |
| Time^2^ | -0.139 | -0.154 – -0.123 | <.001 |
| Sex (male) | -0.105 | -0.226 – 0.015 | .09 |
| *APOE* (*Ɛ2*) | 0.051 | -0.091 – 0.193 | .48 |
| *APOE* (*Ɛ4*) | -0.283 | -0.406 – -0.159 | <.001 |
| Baseline age | -0.449 | -0.493 – -0.405 | <.001 |
| Education | 0.137 | 0.093 – 0.181 | <.001 |
| $\surd$visit number | 0.426 | 0.342 – 0.511 | <.001 |
| Vascular risk | -0.042 | -0.085 – 0.000 | .05 |
| Time | -0.858 | -0.966 – -0.750 | <.001 |
| **Cognition ~ *APOE* × time + [baseline age + education + baseline vascular risk score] × time +** $\boldsymbol{\surd}$**visit number + time^2^; Men; *N* = 471** | | | |
| *APOE* (*Ɛ2*) × time | 0.149 | -0.022 – 0.319 | .09 |
| *APOE* (*Ɛ4*) × time | -0.317 | -0.476 – -0.158 | **<.001** |
| Baseline age × time | -0.235 | -0.298 – -0.171 | **<.001** |
| Education × time | -0.052 | -0.116 – 0.011 | .11 |
| Vascular risk × time | -0.057 | -0.118 – 0.004 | .07 |
| Time^2^ | -0.094 | -0.128 – -0.060 | **<.001** |
| *APOE* (*Ɛ2*) | 0.152 | -0.093 – 0.398 | .22 |
| *APOE* (*Ɛ4*) | -0.433 | -0.658 – -0.208 | **<.001** |
| Baseline age | -0.407 | -0.495 – -0.318 | **<.001** |
| Education | 0.163 | 0.071 – 0.255 | **.001** |
| $\surd$visit number | 0.587 | 0.402 – 0.772 | **<.001** |
| Vascular risk | -0.108 | -0.194 – -0.021 | **.02** |
| Time | -1.035 | -1.264 – -0.806 | **<.001** |
| **Cognition ~ *APOE* × time + [baseline age + education + baseline vascular risk score] × time +** $\boldsymbol{\surd}$**visit number + time^2^; Women; *N* = 1,364** | | | |
| *APOE* (*Ɛ2*) × time | 0.012 | -0.089 – 0.114 | .81 |
| *APOE* (*Ɛ4*) × time | -0.281 | -0.368 – -0.193 | <.001 |
| Baseline age × time | -0.256 | -0.293 – -0.220 | <.001 |
| Education × time | -0.010 | -0.045 – 0.026 | .60 |
| Vascular risk × time | -0.009 | -0.044 – 0.026 | .62 |
| Time^2^ | -0.160 | -0.177 – -0.143 | <.001 |
| *APOE* (*Ɛ2*) | 0.048 | -0.091 – 0.188 | .50 |
| *APOE* (*Ɛ4*) | -0.272 | -0.393 – -0.151 | <.001 |
| Baseline age | -0.455 | -0.505 – -0.406 | <.001 |
| Education | 0.126 | 0.077 – 0.175 | <.001 |
| $\surd$visit number | 0.342 | 0.246 – 0.437 | <.001 |
| Vascular risk | -0.021 | -0.070 – 0.027 | .39 |
| Time | -0.747 | -0.866 – -0.628 | <.001 |
| **Cognition ~ sex × time + [baseline age + education + baseline vascular risk score] × time +** $\boldsymbol{\surd}$**visit number + time^2^; *APOE* *Ɛ2*** **carriers; *N* = 265** | | | |
| Sex (male) × time | 0.191 | 0.012 – 0.371 | **.04** |
| Baseline age × time | -0.240 | -0.325 – -0.154 | **<.001** |
| Education × time | -0.036 | -0.120 – 0.047 | .40 |
| Vascular risk × time | -0.085 | -0.165 – -0.005 | **.04** |
| Time^2^ | -0.078 | -0.121 – -0.035 | **<.001** |
| Sex (male) | -0.009 | -0.262 – 0.244 | .95 |
| Baseline age | -0.389 | -0.506 – -0.272 | **<.001** |
| Education | 0.188 | 0.071 – 0.304 | **.002** |
| $\surd$visit number | 0.596 | 0.363 – 0.830 | **<.001** |
| Vascular risk | -0.084 | -0.196 – 0.029 | .15 |
| Time | -1.082 | -1.368 – -0.795 | **<.001** |
| **Cognition ~ sex × time + [baseline age + education + baseline vascular risk score] × time +** $\boldsymbol{\surd}$**visit number + time^2^; *APOE* *Ɛ3/Ɛ3*** **carriers; *N* = 1,197** | | | |
| Sex (male) × time | 0.038 | -0.044 – 0.121 | .36 |
| Baseline age × time | -0.264 | -0.302 – -0.227 | **<.001** |
| Education × time | -0.020 | -0.057 – 0.017 | .29 |
| Vascular risk × time | 0.005 | -0.031 – 0.040 | .80 |
| Time^2^ | -0.157 | -0.176 – -0.138 | **<.001** |
| Sex (male) | -0.121 | -0.236 – -0.005 | **.04** |
| Baseline age | -0.489 | -0.540 – -0.437 | **<.001** |
| Education | 0.156 | 0.104 – 0.208 | **<.001** |
| $\surd$visit number | 0.360 | 0.254 – 0.466 | **<.001** |
| Vascular risk | -0.021 | -0.071 – 0.029 | .41 |
| Time | -0.795 | -0.925 – -0.664 | **<.001** |

**Table S7. Full regression outputs of linear mixed models examining sex differences in associations between *APOE Ɛ2* (vs. *Ɛ3/Ɛ3*) and longitudinal cognition in non-Hispanic White participants in Sample 2 (ADNI & Prevent-AD).**

|  | Standardized *β* estimate | 95% confidence interval | *P* value |
| --- | --- | --- | --- |
| **Cognition ~ sex × *APOE* × time + [baseline age + education + baseline vascular risk score + cohort] × time + time^2^; *N* = 915** | | | |
| Sex (male) × *APOE* (*Ɛ2*) × time | 0.196 | 0.007 – 0.386 | .04 |
| Sex (male) × *APOE* (*Ɛ4*) × time | 0.041 | -0.095 – 0.177 | .56 |
| Sex (male) × time | -0.004 | -0.086 – 0.078 | .93 |
| *APOE* (*Ɛ2*) × time | -0.108 | -0.240 – 0.023 | .11 |
| *APOE* (*Ɛ4*) × time | -0.159 | -0.247 – -0.071 | <.001 |
| Sex (male) × *APOE* (*Ɛ2*) | 0.254 | -0.082 – 0.589 | .14 |
| Sex (male) × *APOE* (*Ɛ4*) | -0.033 | -0.272 – 0.206 | .79 |
| Baseline age × time | -0.095 | -0.136 – -0.054 | <.001 |
| Education × time | 0.016 | -0.015 – 0.047 | .31 |
| Vascular risk × time | -0.003 | -0.039 – 0.033 | .86 |
| Cohort (Prevent-AD) × time | -0.148 | -0.243 – -0.053 | .002 |
| Time^2^ | -0.090 | -0.105 – -0.076 | <.001 |
| Sex (male) | -0.382 | -0.527 – -0.236 | <.001 |
| *APOE* (*Ɛ2*) | -0.102 | -0.333 – 0.128 | .38 |
| *APOE* (*Ɛ4*) | -0.075 | -0.225 – 0.075 | .33 |
| Baseline age | -0.350 | -0.420 – -0.280 | <.001 |
| Education | 0.246 | 0.191 – 0.302 | <.001 |
| Vascular risk | -0.023 | -0.083 – 0.038 | .46 |
| Cohort (Prevent-AD) | -0.493 | -0.655 – -0.332 | <.001 |
| Time | 0.046 | -0.017 – 0.110 | .15 |
| **Cognition ~ *APOE* × time + [baseline age + education + baseline vascular risk score + cohort] × time + time^2^; Men; *N* = 373** | | | |
| *APOE* (*Ɛ2*) × time | 0.094 | -0.055 – 0.244 | .22 |
| *APOE* (*Ɛ4*) × time | -0.133 | -0.250 – -0.017 | **.03** |
| Baseline age × time | -0.086 | -0.152 – -0.021 | **.01** |
| Education × time | -0.008 | -0.060 – 0.045 | .77 |
| Vascular risk × time | -0.001 | -0.057 – 0.056 | .99 |
| Cohort (Prevent-AD) × time | -0.114 | -0.282 – 0.054 | .18 |
| Time^2^ | -0.086 | -0.109 – -0.063 | **<.001** |
| *APOE* (*Ɛ2*) | 0.160 | -0.096 – 0.417 | .22 |
| *APOE* (*Ɛ4*) | -0.121 | -0.320 – 0.077 | .23 |
| Baseline age | -0.330 | -0.439 – -0.221 | **<.001** |
| Education | 0.191 | 0.102 – 0.280 | **<.001** |
| Vascular risk | -0.055 | -0.150 – 0.040 | .26 |
| Cohort (Prevent-AD) | -0.498 | -0.784 – -0.213 | **.001** |
| Time | -0.008 | -0.084 – 0.067 | .83 |
| **Cognition ~ *APOE* × time + [baseline age + education + baseline vascular risk score + cohort] × time + time^2^; Women; *N* = 542** | | | |
| *APOE* (*Ɛ2*) × time | -0.103 | -0.227 – 0.020 | .10 |
| *APOE* (*Ɛ4*) × time | -0.151 | -0.234 – -0.068 | **<.001** |
| Baseline age × time | -0.088 | -0.139 – -0.037 | **.001** |
| Education × time | 0.026 | -0.011 – 0.063 | .17 |
| Vascular risk × time | -0.004 | -0.051 – 0.042 | .85 |
| Cohort (Prevent-AD) × time | -0.158 | -0.272 – -0.044 | **.007** |
| Time^2^ | -0.098 | -0.117 – -0.080 | **<.001** |
| *APOE* (*Ɛ2*) | -0.094 | -0.322 – 0.133 | .42 |
| *APOE* (*Ɛ4*) | -0.068 | -0.217 – 0.081 | .37 |
| Baseline age | -0.354 | -0.446 – -0.263 | **<.001** |
| Education | 0.271 | 0.201 – 0.340 | **<.001** |
| Vascular risk | 0.001 | -0.079 – 0.080 | .99 |
| Cohort (Prevent-AD) | -0.488 | -0.686 – -0.289 | **<.001** |
| Time | 0.083 | 0.012 – 0.154 | **.02** |
| **Cognition ~ sex × time + [baseline age + education + baseline vascular risk score + cohort] × time + time^2^; *APOE* *Ɛ2*** **carriers; *N* = 108** | | | |
| Sex (male) × time | 0.151 | -0.008 – 0.311 | .06 |
| Baseline age × time | -0.014 | -0.103 – 0.075 | .76 |
| Education × time | 0.023 | -0.053 – 0.099 | .56 |
| Vascular risk × time | -0.018 | -0.112 – 0.076 | .71 |
| Cohort (Prevent-AD) × time | -0.047 | -0.267 – 0.174 | .68 |
| Time^2^ | -0.077 | -0.123 – -0.032 | **.001** |
| Sex (male) | -0.218 | -0.554 – 0.117 | .20 |
| Baseline age | -0.282 | -0.476 – -0.089 | **.004** |
| Education | 0.323 | 0.149 – 0.498 | **<.001** |
| Vascular risk | -0.027 | -0.214 – 0.161 | .78 |
| Cohort (Prevent-AD) | -0.425 | -0.890 – 0.040 | .07 |
| Time | -0.092 | -0.227 – 0.044 | .19 |
| **Cognition ~ sex × time + [baseline age + education + baseline vascular risk score + cohort] × time + time^2^; *APOE* *Ɛ3/Ɛ3*** **carriers; *N* = 520** | | | |
| Sex (male) × time | -0.005 | -0.088 – 0.077 | .90 |
| Baseline age × time | -0.095 | -0.147 – -0.043 | **<.001** |
| Education × time | 0.014 | -0.026 – 0.053 | .51 |
| Vascular risk × time | -0.000 | -0.046 – 0.045 | >.99 |
| Cohort (Prevent-AD) × time | -0.142 | -0.264 – -0.020 | **.02** |
| Time^2^ | -0.089 | -0.108 – -0.070 | **<.001** |
| Sex (male) | -0.401 | -0.549 – -0.254 | **<.001** |
| Baseline age | -0.342 | -0.431 – -0.252 | **<.001** |
| Education | 0.249 | 0.178 – 0.320 | **<.001** |
| Vascular risk | -0.044 | -0.121 – 0.033 | .26 |
| Cohort (Prevent-AD) | -0.556 | -0.766 – -0.345 | **<.001** |
| Time | 0.027 | -0.042 – 0.095 | .45 |
| **Cognition ~ sex × time + [baseline age + education + baseline vascular risk score + cohort] × time + time^2^; *APOE* *Ɛ4*** **carriers; *N* = 287** | | | |
| Sex (male) × time | 0.031 | -0.091 – 0.153 | .62 |
| Baseline age × time | -0.138 | -0.222 – -0.053 | **.001** |
| Education × time | 0.012 | -0.049 – 0.072 | .71 |
| Vascular risk × time | -0.008 | -0.080 – 0.064 | .83 |
| Cohort (Prevent-AD) × time | -0.255 | -0.447 – -0.062 | **.009** |
| Time^2^ | -0.105 | -0.130 – -0.080 | **<.001** |
| Sex (male) | -0.385 | -0.586 – -0.183 | **<.001** |
| Baseline age | -0.391 | -0.527 – -0.256 | **<.001** |
| Education | 0.222 | 0.119 – 0.326 | **<.001** |
| Vascular risk | 0.014 | -0.101 – 0.129 | .81 |
| Cohort (Prevent-AD) | -0.461 | -0.765 – -0.157 | **.003** |
| Time | -0.019 | -0.130 – 0.093 | .75 |

**Table S8. Full regression outputs of linear mixed models examining sex differences in associations between *APOE Ɛ2* (vs. *Ɛ3/Ɛ3*) and longitudinal cognition in non-Hispanic White participants in Sample 1 (NACC & ROS/MAP) at increasing baseline age cut-offs.**

|  | Standardized *β* estimate | 95% confidence interval | *P* value |
| --- | --- | --- | --- |
| **Cognition ~ sex × *APOE* × time + [baseline age + education + baseline vascular risk score + cohort] × time +** $\boldsymbol{\surd}$**visit number + time^2^, Baseline age ≥ 65, *N* = 8,112** | | | |
| Sex (male) × *APOE* (*Ɛ2*) × time | 0.097 | 0.012 – 0.182 | .03 |
| Sex (male) × *APOE* (*Ɛ4*) × time | 0.080 | 0.014 – 0.147 | .02 |
| Sex (male) × time | 0.003 | -0.034 – 0.039 | .88 |
| *APOE* (*Ɛ2*) × time | 0.003 | -0.046 – 0.052 | .92 |
| *APOE* (*Ɛ4*) × time | -0.225 | -0.265 – -0.185 | <.001 |
| Sex (male) × *APOE* (*Ɛ2*) | 0.143 | 0.020 – 0.266 | .02 |
| Sex (male) × *APOE* (*Ɛ4*) | 0.070 | -0.025 – 0.164 | .15 |
| Baseline age × time | -0.195 | -0.208 – -0.181 | <.001 |
| Education × time | 0.003 | -0.011 – 0.017 | .63 |
| Vascular risk × time | -0.006 | -0.021 – 0.008 | .40 |
| Cohort (ROS/MAP) × time | 0.051 | 0.017 – 0.084 | .003 |
| Time^2^ | -0.140 | -0.146 – -0.135 | <.001 |
| Sex (male) | -0.195 | -0.248 – -0.143 | <.001 |
| *APOE* (*Ɛ2*) | -0.003 | -0.074 – 0.069 | .95 |
| *APOE* (*Ɛ4*) | -0.291 | -0.348 – -0.234 | <.001 |
| Baseline age | -0.395 | -0.415 – -0.376 | <.001 |
| Education | 0.224 | 0.204 – 0.245 | <.001 |
| $\surd$visit number | 0.104 | 0.084 – 0.123 | <.001 |
| Vascular risk | -0.036 | -0.057 – -0.015 | .001 |
| Cohort (ROS/MAP) | -0.135 | -0.185 – -0.086 | <.001 |
| Time | -0.345 | -0.379 – -0.310 | <.001 |
| **Cognition ~ sex × *APOE* × time + [baseline age + education + baseline vascular risk score + cohort] × time +** $\boldsymbol{\surd}$**visit number + time^2^, Baseline age ≥ 70, *N* = 6,206** | | | |
| Sex (male) × *APOE* (*Ɛ2*) × time | 0.086 | -0.009 – 0.181 | .08 |
| Sex (male) × *APOE* (*Ɛ4*) × time | 0.081 | 0.003 – 0.159 | .04 |
| Sex (male) × time | 0.005 | -0.036 – 0.046 | .81 |
| *APOE* (*Ɛ2*) × time | 0.025 | -0.030 – 0.079 | .38 |
| *APOE* (*Ɛ4*) × time | -0.222 | -0.268 – -0.175 | <.001 |
| Sex (male) × *APOE* (*Ɛ2*) | 0.149 | 0.010 – 0.288 | .04 |
| Sex (male) × *APOE* (*Ɛ4*) | 0.063 | -0.051 – 0.177 | .28 |
| Baseline age × time | -0.155 | -0.170 – -0.139 | <.001 |
| Education × time | 0.009 | -0.006 – 0.025 | .24 |
| Vascular risk × time | -0.006 | -0.023 – 0.011 | .48 |
| Cohort (ROS/MAP) × time | 0.028 | -0.009 – 0.066 | .14 |
| Time^2^ | -0.149 | -0.155 – -0.142 | <.001 |
| Sex (male) | -0.192 | -0.252 – -0.131 | <.001 |
| *APOE* (*Ɛ2*) | 0.028 | -0.054 – 0.110 | .50 |
| *APOE* (*Ɛ4*) | -0.309 | -0.378 – -0.241 | <.001 |
| Baseline age | -0.335 | -0.358 – -0.313 | <.001 |
| Education | 0.225 | 0.202 – 0.249 | <.001 |
| $\surd$visit number | 0.063 | 0.040 – 0.086 | <.001 |
| Vascular risk | -0.036 | -0.060 – -0.011 | .004 |
| Cohort (ROS/MAP) | -0.135 | -0.191 – -0.079 | <.001 |
| Time | -0.335 | -0.374 – -0.295 | <.001 |
| **Cognition ~ sex × *APOE* × time + [baseline age + education + baseline vascular risk score + cohort] × time +** $\boldsymbol{\surd}$**visit number + time^2^, Baseline age ≥ 75, *N* = 4,156** | | | |
| Sex (male) × *APOE* (*Ɛ2*) × time | 0.103 | -0.007 – 0.214 | .07 |
| Sex (male) × *APOE* (*Ɛ4*) × time | 0.087 | -0.012 – 0.185 | .08 |
| Sex (male) × time | -0.008 | -0.058 – 0.041 | .74 |
| *APOE* (*Ɛ2*) × time | 0.031 | -0.031 – 0.094 | .32 |
| *APOE* (*Ɛ4*) × time | -0.205 | -0.264 – -0.147 | <.001 |
| Sex (male) × *APOE* (*Ɛ2*) | 0.156 | -0.010 – 0.322 | .07 |
| Sex (male) × *APOE* (*Ɛ4*) | 0.011 | -0.136 – 0.158 | .88 |
| Baseline age × time | -0.127 | -0.146 – -0.109 | <.001 |
| Education × time | 0.015 | -0.004 – 0.034 | .13 |
| Vascular risk × time | -0.001 | -0.021 – 0.019 | .90 |
| Cohort (ROS/MAP) × time | 0.020 | -0.023 – 0.064 | .36 |
| Time^2^ | -0.157 | -0.164 – -0.149 | <.001 |
| Sex (male) | -0.177 | -0.251 – -0.104 | <.001 |
| *APOE* (*Ɛ2*) | 0.049 | -0.047 – 0.145 | .32 |
| *APOE* (*Ɛ4*) | -0.277 | -0.366 – -0.189 | <.001 |
| Baseline age | -0.273 | -0.300 – -0.247 | <.001 |
| Education | 0.238 | 0.209 – 0.267 | <.001 |
| $\surd$visit number | 0.029 | 0.000 – 0.058 | .05 |
| Vascular risk | -0.032 | -0.062 – -0.002 | .04 |
| Cohort (ROS/MAP) | -0.092 | -0.158 – -0.026 | .007 |
| Time | -0.337 | -0.386 – -0.288 | <.001 |
| **Cognition ~ sex × *APOE* × time + [baseline age + education + baseline vascular risk score + cohort] × time +** $\boldsymbol{\surd}$**visit number + time^2^, Baseline age ≥ 80, *N* = 2,419** | | | |
| Sex (male) × *APOE* (*Ɛ2*) × time | 0.134 | -0.006 – 0.274 | .06 |
| Sex (male) × *APOE* (*Ɛ4*) × time | 0.059 | -0.071 – 0.190 | .37 |
| Sex (male) × time | 0.008 | -0.057 – 0.072 | .82 |
| *APOE* (*Ɛ2*) × time | 0.012 | -0.068 – 0.092 | .77 |
| *APOE* (*Ɛ4*) × time | -0.185 | -0.262 – -0.107 | <.001 |
| Sex (male) × *APOE* (*Ɛ2*) | 0.256 | 0.041 – 0.471 | .02 |
| Sex (male) × *APOE* (*Ɛ4*) | 0.052 | -0.147 – 0.251 | .61 |
| Baseline age × time | -0.106 | -0.130 – -0.082 | <.001 |
| Education × time | 0.009 | -0.016 – 0.034 | .48 |
| Vascular risk × time | 0.013 | -0.013 – 0.038 | .34 |
| Cohort (ROS/MAP) × time | -0.002 | -0.059 – 0.054 | .94 |
| Time^2^ | -0.152 | -0.162 – -0.142 | <.001 |
| Sex (male) | -0.175 | -0.273 – -0.078 | <.001 |
| *APOE* (*Ɛ2*) | 0.014 | -0.110 – 0.138 | .82 |
| *APOE* (*Ɛ4*) | -0.271 | -0.390 – -0.151 | <.001 |
| Baseline age | -0.229 | -0.265 – -0.194 | <.001 |
| Education | 0.245 | 0.207 – 0.283 | <.001 |
| $\surd$visit number | 0.014 | -0.024 – 0.052 | .47 |
| Vascular risk | -0.011 | -0.051 – 0.028 | .57 |
| Cohort (ROS/MAP) | -0.082 | -0.168 – 0.005 | .06 |
| Time | -0.346 | -0.410 – -0.282 | <.001 |

**Table S9. Full regression outputs of linear mixed models examining sex differences in associations between *APOE Ɛ2* (vs. *Ɛ3/Ɛ3*) and longitudinal cognition in non-Hispanic White participants in Sample 2 (ADNI & Prevent-AD) at increasing baseline age cut-offs.**

|  | Standardized *β* estimate | 95% confidence interval | *P* value |
| --- | --- | --- | --- |
| **Cognition ~ sex × *APOE* × time + [baseline age + education + baseline vascular risk score + cohort] × time + time^2^; Baseline age ≥ 65, *N* = 684** | | | |
| Sex (male) × *APOE* (*Ɛ2*) × time | 0.210 | -0.016 – 0.435 | .07 |
| Sex (male) × *APOE* (*Ɛ4*) × time | 0.063 | -0.105 – 0.231 | .46 |
| Sex (male) × time | -0.025 | -0.120 – 0.071 | .62 |
| *APOE* (*Ɛ2*) × time | -0.120 | -0.282 – 0.043 | .15 |
| *APOE* (*Ɛ4*) × time | -0.175 | -0.289 – -0.061 | **.003** |
| Sex (male) × *APOE* (*Ɛ2*) | 0.216 | -0.174 – 0.606 | .28 |
| Sex (male) × *APOE* (*Ɛ4*) | -0.026 | -0.311 – 0.258 | .86 |
| Baseline age × time | -0.080 | -0.118 – -0.041 | **<.001** |
| Education × time | 0.027 | -0.011 – 0.065 | .17 |
| Vascular risk × time | -0.017 | -0.058 – 0.024 | .42 |
| Cohort (Prevent-AD) × time | -0.156 | -0.275 – -0.037 | **.01** |
| Time^2^ | -0.098 | -0.114 – -0.082 | **<.001** |
| Sex (male) | -0.357 | -0.521 – -0.192 | **<.001** |
| *APOE* (*Ɛ2*) | -0.058 | -0.342 – 0.225 | .69 |
| *APOE* (*Ɛ4*) | -0.113 | -0.300 – 0.075 | .24 |
| Baseline age | -0.288 | -0.351 – -0.224 | **<.001** |
| Education | 0.287 | 0.221 – 0.354 | **<.001** |
| Vascular risk | -0.026 | -0.093 – 0.040 | .44 |
| Cohort (Prevent-AD) | -0.345 | -0.546 – -0.144 | **.001** |
| Time | 0.008 | -0.061 – 0.076 | .83 |
| **Cognition ~ sex × *APOE* × time + [baseline age + education + baseline vascular risk score + cohort] × time + time^2^; Baseline age ≥ 70, *N* = 464** | | | |
| Sex (male) × *APOE* (*Ɛ2*) × time | 0.219 | -0.062 – 0.499 | .13 |
| Sex (male) × *APOE* (*Ɛ4*) × time | 0.250 | 0.029 – 0.471 | **.03** |
| Sex (male) × time | -0.035 | -0.154 – 0.085 | .57 |
| *APOE* (*Ɛ2*) × time | -0.098 | -0.307 – 0.111 | .36 |
| *APOE* (*Ɛ4*) × time | -0.305 | -0.461 – -0.148 | **<.001** |
| Sex (male) × *APOE* (*Ɛ2*) | 0.427 | -0.053 – 0.907 | .08 |
| Sex (male) × *APOE* (*Ɛ4*) | 0.092 | -0.283 – 0.467 | .63 |
| Baseline age × time | -0.038 | -0.086 – 0.009 | .11 |
| Education × time | 0.032 | -0.018 – 0.081 | .21 |
| Vascular risk × time | -0.010 | -0.060 – 0.040 | .69 |
| Cohort (Prevent-AD) × time | -0.024 | -0.204 – 0.157 | .80 |
| Time^2^ | -0.108 | -0.127 – -0.089 | **<.001** |
| Sex (male) | -0.409 | -0.613 – -0.205 | **<.001** |
| *APOE* (*Ɛ2*) | -0.169 | -0.527 – 0.189 | .36 |
| *APOE* (*Ɛ4*) | -0.245 | -0.503 – 0.013 | .06 |
| Baseline age | -0.143 | -0.223 – -0.064 | **<.001** |
| Education | 0.247 | 0.162 – 0.332 | **<.001** |
| Vascular risk | -0.001 | -0.084 – 0.082 | .98 |
| Cohort (Prevent-AD) | -0.342 | -0.652 – -0.033 | **.03** |
| Time | -0.055 | -0.141 – 0.030 | .21 |
| **Cognition ~ sex × *APOE* × time + [baseline age + education + baseline vascular risk score + cohort] × time + time^2^; Baseline age ≥ 75, *N* = 245** | | | |
| Sex (male) × *APOE* (*Ɛ2*) × time | 0.143 | -0.216 – 0.502 | .44 |
| Sex (male) × *APOE* (*Ɛ4*) × time | 0.116 | -0.177 – 0.410 | .44 |
| Sex (male) × time | -0.053 | -0.207 – 0.102 | .50 |
| *APOE* (*Ɛ2*) × time | 0.075 | -0.190 – 0.340 | .58 |
| *APOE* (*Ɛ4*) × time | -0.187 | -0.394 – 0.021 | .08 |
| Sex (male) × *APOE* (*Ɛ2*) | 0.347 | -0.335 – 1.029 | .32 |
| Sex (male) × *APOE* (*Ɛ4*) | 0.163 | -0.360 – 0.685 | .54 |
| Baseline age × time | 0.008 | -0.054 – 0.070 | .79 |
| Education × time | 0.030 | -0.035 – 0.094 | .36 |
| Vascular risk × time | 0.002 | -0.062 – 0.065 | .96 |
| Cohort (Prevent-AD) × time | -0.125 | -0.444 – 0.194 | .44 |
| Time^2^ | -0.096 | -0.120 – -0.072 | **<.001** |
| Sex (male) | -0.404 | -0.680 – -0.127 | **.004** |
| *APOE* (*Ɛ2*) | -0.076 | -0.572 – 0.420 | .76 |
| *APOE* (*Ɛ4*) | -0.414 | -0.786 – -0.042 | **.03** |
| Baseline age | -0.123 | -0.231 – -0.015 | **.03** |
| Education | 0.275 | 0.156 – 0.394 | **<.001** |
| Vascular risk | -0.024 | -0.134 – 0.086 | .67 |
| Cohort (Prevent-AD) | -0.509 | -1.135 – 0.117 | .11 |
| Time | -0.108 | -0.214 – -0.001 | **.05** |
| **Cognition ~ sex × *APOE* × time + [baseline age + education + baseline vascular risk score + cohort] × time + time^2^; Baseline age ≥ 80, *N* = 88** | | | |
| Sex (male) × *APOE* (*Ɛ2*) × time | 0.044 | -0.563 – 0.652 | .89 |
| Sex (male) × *APOE* (*Ɛ4*) × time | 0.458 | -0.040 – 0.956 | .07 |
| Sex (male) × time | -0.138 | -0.393 – 0.117 | .29 |
| *APOE* (*Ɛ2*) × time | 0.181 | -0.339 – 0.700 | .50 |
| *APOE* (*Ɛ4*) × time | -0.430 | -0.842 – -0.019 | **.04** |
| Sex (male) × *APOE* (*Ɛ2*) | -0.169 | -1.337 – 0.999 | .78 |
| Sex (male) × *APOE* (*Ɛ4*) | 0.616 | -0.374 – 1.606 | .22 |
| Baseline age × time | 0.068 | -0.031 – 0.167 | .18 |
| Education × time | 0.013 | -0.097 – 0.123 | .82 |
| Vascular risk × time | -0.041 | -0.136 – 0.054 | .40 |
| Cohort (Prevent-AD) × time | -0.107 | -0.670 – 0.456 | .71 |
| Time^2^ | -0.118 | -0.163 – -0.073 | **<.001** |
| Sex (male) | -0.826 | -1.296 – -0.357 | **.001** |
| *APOE* (*Ɛ2*) | 0.299 | -0.635 – 1.233 | .53 |
| *APOE* (*Ɛ4*) | -0.548 | -1.379 – 0.283 | .20 |
| Baseline age | 0.053 | -0.127 – 0.232 | .57 |
| Education | 0.362 | 0.157 – 0.567 | **.001** |
| Vascular risk | -0.066 | -0.243 – 0.112 | .47 |
| Cohort (Prevent-AD) | -1.038 | -2.263 – 0.187 | .10 |
| Time | -0.031 | -0.214 – 0.153 | .75 |

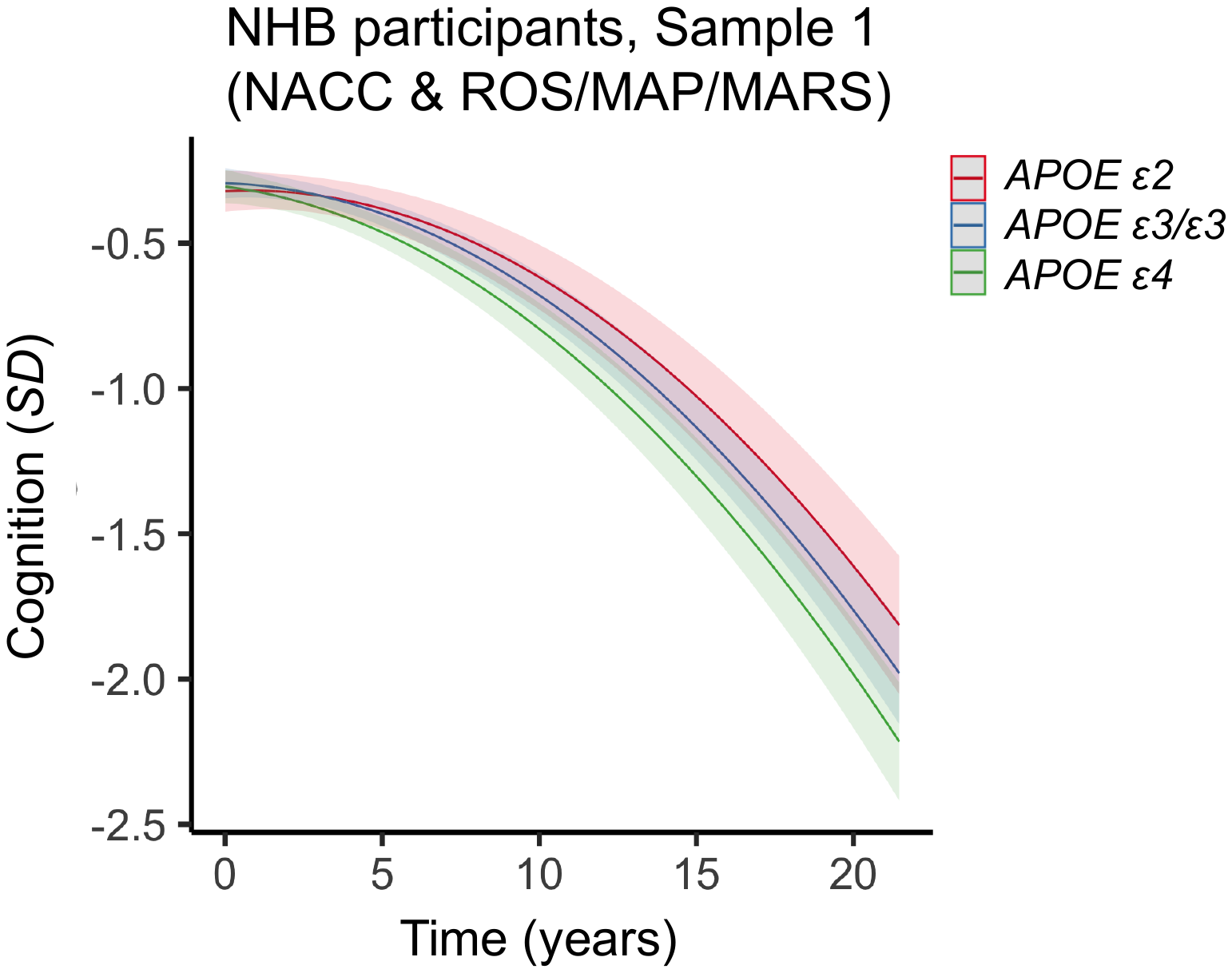

**Figure S2.** **Two-way interaction between *APOE* and time on cognitive decline in non-Hispanic Black (NHB) participants in Sample 1 (NACC & ROS/MAP/MARS).** Plots depict marginal effects, showing change in cognition (standardized score) over time by genotype. The models are adjusted for sex, baseline age, years of education, and vascular risk, and their interactions with time. Shaded regions represent 95% confidence intervals. In NHB participants, *APOE ε2* carriers exhibit a pattern of slower cognitive decline over time relative to *APOE ε3/ε3* carriers, but the interaction with time was not significant.

**Figure S3.** **Two-way interaction between sex and time on cognitive decline in non-Hispanic Black (NHB) *APOE ε4* carriers** **in pooled NACC and ROS/MAP/MARS cohort.** Plots depict marginal effects, showing change in cognition (standardized score) over time by sex. The models are adjusted for sex, baseline age, years of education, and vascular risk, and their interactions with time. Shaded regions represent 95% confidence intervals. Among NHB *APOE ε4* carriers, women exhibit a trend towards faster cognitive decline compared to men.

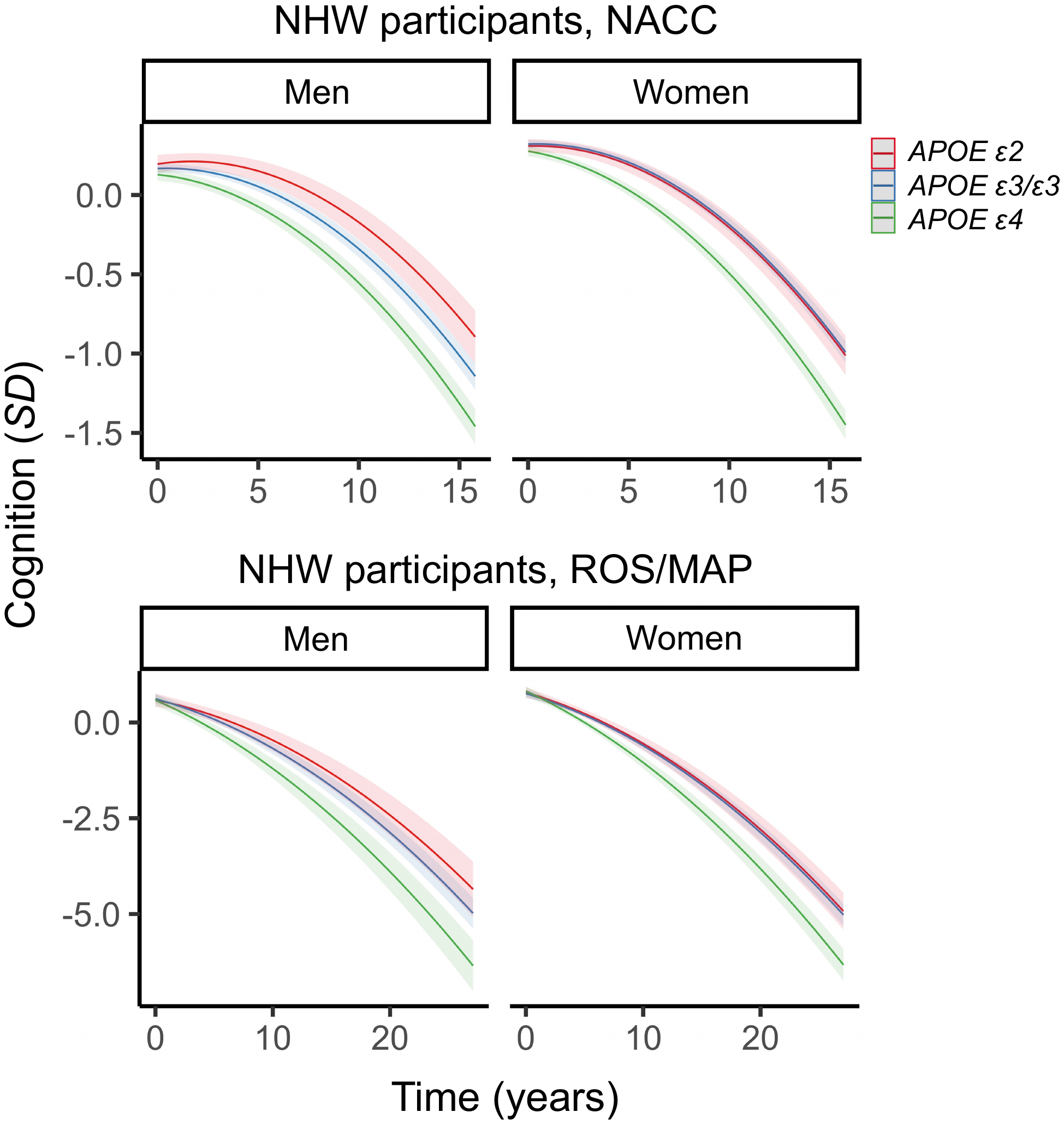

**Figure S4. Three-way interactions between sex, *APOE* and time on cognitive decline among non-Hispanic White (NHW) participants each in NACC and ROS/MAP.** Plots depict marginal effects, showing change in cognition (standardized score) over time in each cohort, stratified by sex. The models are adjusted for baseline age, years of education, and vascular risk, and their interactions with time. Shaded regions represent 95% CIs. Across both cohorts, there is a similar pattern of cognitive protection in men carrying *APOE ε2* (vs. *ε3/ε3*), but no difference in women carrying *APOE ε2* (vs. *ε3/ε3*).

**Figure S5. Three-way interactions between sex, *APOE* and time on cognitive decline among non-Hispanic White (NHW) participants each in NACC and ROS/MAP.** Plots depict marginal effects, showing change in cognition (standardized score) over time in each cohort, stratified by genotype (*APOE ε4* not shown). The models are adjusted for baseline age, years of education, and vascular risk, and their interactions with time. Shaded regions represent 95% CIs. Across both cohorts, there is a similar pattern of cognitive protection in men (vs. women) carrying *APOE ε2.*

**Figure S6.** **Two-way interaction between sex and time on cognitive decline in non-Hispanic White (NHW) *APOE ε4* carriers** **in pooled NACC and ROS/MAP cohort.** Plots depict marginal effects, showing change in cognition (standardized score) over time by sex. The models are adjusted for sex, baseline age, years of education, and vascular risk, and their interactions with time. Shaded regions represent 95% confidence intervals. Among NHW *APOE ε4* carriers, women have significantly faster cognitive decline than men.
